# Associations of Prenatal Cannabis Exposure and Neonatal Brain Development in the HBCD Cohort

**DOI:** 10.64898/2026.03.02.26347436

**Authors:** Leela Shah, Elizabeth M. Planalp, Ryan McDonald, Caitlin J Regner, Sreevalli Atluru, Stephanie L. Merhar, Andrew L. Alexander, Pilar N. Ossorio, Julie Poehlmann, Douglas C. Dean

**Affiliations:** Waisman Center, University of Wisconsin–Madison, 1500 Highland Ave, Madison, WI 53705, USA; Neuroscience Training Program, University of Wisconsin–Madison, 1111 Highland Ave, Madison, WI 53705, USA; Department of Obstetrics and Gynecology, University of Wisconsin–Madison, 600 Highland Ave, Madison, WI 53792, USA; Department of Family Medicine and Community Health, University of Wisconsin–Madison, 600 Highland Ave, Madison, WI 53792, USA; Department of Pediatrics, University of Cincinnati, 3333 Burnet Avenue, Cincinnati, OH 45229; Perinatal Institute, Cincinnati Children’s Hospital Medical Center, 3333 Burnet Avenue, Cincinnati, OH 45229; Department of Medical Physics, University of Wisconsin–Madison, 600 Highland Ave, Madison, WI 53792, USA.; Department of Psychiatry, University of Wisconsin–Madison, 600 Highland Ave, Madison, WI 53792, USA; School of Law, University of Wisconsin–Madison, 600 Highland Ave, Madison, WI 53792, USA; Morgridge Institute for Research, University of Wisconsin–Madison, 600 Highland Ave, Madison, WI 53792, USA; Department of Medical History and Bioethics, University of Wisconsin–Madison, 600 Highland Ave, Madison, WI 53792, USA; Department of Human Development and Family Studies, University of Wisconsin–Madison, 600 Highland Ave, Madison, WI 53792, USA; Department of Pediatrics, University of Wisconsin–Madison, 600 Highland Ave, Madison, WI 53792, USA

**Keywords:** Prenatal cannabis exposure, Neonatal brain development, Diffusion magnetic resonance imaging, White matter microstructure, Prenatal nicotine exposure, HEALthy Brain and Child Development (HBCD) Study

## Abstract

Prenatal cannabis exposure (PCE) is increasing in prevalence, yet its effects on brain development remain incompletely understood. We examined associations between maternal PCE and neonatal brain volumes and white matter microstructure and secondarily assessed the influence of prenatal nicotine exposure (PNE). This cohort study included 1,630 mother–infant dyads (222 with PCE) from the HEALthy Brain and Child Development Study. Maternal substance use was assessed using the Timeline Follow-Back method. Infants underwent natural-sleep T2-weighted structural and diffusion MRI within the first month of life. Analyses evaluated associations by exposure presence, gestational timing, and frequency among exposed infants. Secondary analyses examined PNE as a predictor and tested cannabis × nicotine interactions. PCE was associated with frequency-dependent increases in diffusivity values across white matter regions that remained significant after accounting for PNE. Timing-specific associations emerged, with first-trimester exposure linked to higher diffusivity values within cerebellar white matter regions and sustained exposure throughout pregnancy associated with higher diffusivity values within white matter association regions. In contrast, associations with subcortical brain volumes were attenuated after accounting for PNE, and significant PCE × PNE interactions across several subcortical regions suggested greater sensitivity of volumetric measures to nicotine co-exposure. Effect sizes were small for PCE presence and timing analyses and medium-to-large for PCE frequency analyses. These findings may reflect variation in early neurodevelopment rather than uniformly adverse effects and warrant longitudinal follow-up to determine their relationship with later clinical outcomes. Consideration of exposure timing, frequency, and co-occurring substance use may improve individualized counseling and developmental monitoring.

## Introduction

As cannabis availability has increased across the United States, use has increased ^1^, including in pregnant individuals among whom prenatal cannabis use is estimated at 8% ^2^. Cannabis may be perceived as a natural remedy for pregnancy-related symptoms and anxiety, driving use in this population ^3^. Beyond perinatal outcomes including low birth weight, prematurity, and placental pathologies ^4^, prenatal cannabis exposure (PCE) is associated with neurodevelopmental sequelae, including memory deficits, impulsivity, hyperactivity, and addiction vulnerability ^5,6^. Yet, the neurobiological mechanisms underlying PCE-related outcomes remain incompletely understood.

Cannabis can disrupt the developing endocannabinoid system, which regulates processes including neuronal migration, axon guidance, synaptogenesis, circuit refinement, and oligodendrocyte development ^6–10^. The endocannabinoid system undergoes changes throughout gestation, exhibiting region- and timing-specific patterns of ligand and receptor expression ^11,12^. Endocannabinoid signaling is tightly regulated during fetal brain development, and cannabinoid receptor expression is detectable by 5–6 weeks of gestation in the prefrontal cortex, mesolimbic system, striatum, cerebellum, and hypothalamic–pituitary axis ^6,7,13^. Because endocannabinoid receptor expression, signaling, and metabolic pathways change across gestation, PCE may differentially affect neurodevelopment depending on the timing, frequency, and persistence of exposure ^14^. Furthermore, PCE frequently co-occurs with exposure to other substances, most commonly nicotine, which may lead to cumulative or synergistic effects on neurodevelopment ^15^ However, animal and cellular models cannot fully replicate the pharmacokinetics of PCE in humans, limiting translation of existing work ^16,17^.

Human neuroimaging studies have shown associations between PCE and cerebellar ^18^, cortical ^18–20^, fronto- and meso-limbic (e.g., the insula and fornix) ^18,19,21^, white matter relay (e.g., the internal capsule) ^19,21,22^, and basal ganglia ^18^ development. Yet, most studies are conducted during adolescence, when retrospective reports, continued postnatal exposures, and other experiences may affect the measured association between PCE and neurodevelopment ^20–22^. Critically, PCE continuing beyond early gestation may confer greater risk for adverse neurodevelopmental and behavioral outcomes ^19,21,23^, and early differences may be amplified and shape later development ^24^. These findings underscore the need to examine timing- and frequency-dependent associations of PCE and neurodevelopment within large samples. Infant neuroimaging work further indicates regionally specific alterations in brain growth trajectories, with accelerated growth in rapidly developing regions and slowed growth in other regions that appears dose dependent ^19^. However, existing infant neuroimaging studies of PCE have small sample sizes and limited ability to model clinically relevant patterns of exposure or broader contextual sociodemographic and psychosocial influences ^25^.

This study analyzes data from the HEALthy Brain and Child Development (HBCD) Study, the largest longitudinal study of brain development in the U.S. ^26,27^. HBCD aims to address challenges in studying prenatal substance exposure, including stigma, legal concerns, and underreporting ^28,29^. Its trauma-informed approach supports accurate disclosure and retention across diverse populations ^30,31^. We examined associations between patterns of maternal PCE, including presence, gestational timing, and frequency, and measures of neonatal gray matter structure (T2-weighted regional brain volumes) and white matter microstructure (diffusion tensor imaging metrics), while accounting for sociodemographic factors. Exploratory analyses examined associations between PCE, PNE, and their interaction and brain metrics ^15,32^.

## Methods

### Study Design and Participants

This observational cohort study analyzed data from the HBCD Study Data Release 2.0 ^33,34^. Of 2,310 available dyads, 1,857 had volumetric data. 227 had sub-threshold PCE and were excluded, leading to 1,630 dyads in the sample (222 with PCE, eFigure 1 in the Supplement). Diffusion MRI data were available for 1,242 dyads (176 with PCE). Pregnant participants completed prenatal assessments during the second or third trimester. Infants underwent MRI in the first month of life using harmonized multisite protocols ^26^. Written informed consent was obtained from all participants, and study procedures were approved by the Institutional Review Board (IRB) of record at the University of California, San Diego, and by local IRBs at HBCD implementation sites. Cohort and imaging methods are described elsewhere ^26,27^. This study followed the Strengthening the Reporting of Observational Studies in Epidemiology (STROBE) reporting guideline ^35^.

### PCE Quantification and Covariates

Prenatal substance exposure was assessed through maternal self-report using the validated HBCD Modified ASSIST ^36^ instrument, which screened for substance use during pregnancy. Participants who screened positive completed the Timeline Follow Back assessment (TLFB, eFigure 2 in the Supplement) ^37–39^, which captured the number of uses per week across 7 sampled weeks spanning gestation. PCE was defined as any maternal prenatal cannabis use during at least 4 of the 7 TLFB-assessed weeks ^40^. Based on HBCD analytic workflows ^40–42^, this threshold was selected a priori to distinguish sustained exposure from isolated or sporadic exposure. Participants with sub-threshold PCE were excluded from analyses. Within dyads meeting the PCE threshold with sufficient TLFB data to assign a timing category, first trimester PCE was defined as PCE from 2-6 weeks post-conception (TLFB weeks 1–4, eFigure 2 in the Supplement). Sustained PCE was defined as first trimester PCE combined with use during the week of the prenatal study visit (second or third trimester) or the 2 weeks preceding delivery (TLFB weeks 5–7, eFigure 2 in the Supplement). Within subjects with TLFB data available for all 7 weeks, frequency of exposure was defined as the total number of cannabis use episodes across the 7 TLFB-assessed weeks. Maternal mental health symptoms were assessed using the Edinburgh Postnatal Depression Scale (EPDS) ^43^ and the Diagnostic and Statistical Manual of Mental Disorders, 5^th^ Edition (DSM-5) Self-Rated Cross-Cutting Anxiety Measure Level 1 ^44^. Additional covariates included maternal age, educational attainment, household income, and maternal prenatal nicotine (≥ 4 weeks), alcohol (≥ 4 weeks), or opioid (≥ 2 weeks) exposure. Infant sex and corrected gestational age at MRI were included in all models.

### Neuroimaging Acquisition, Processing, and Harmonization

Neonatal MRI data were acquired using harmonized HBCD infant protocols optimized for natural sleep ^26^. This analysis focused on T2-weighted structural MRI and diffusion MRI (dMRI). T2-weighted contrasts were used for segmentation, quality control, and analysis as they provide superior gray-white matter contrast in the neonatal brain compared to T1-weighted contrasts ^45,46^. Moreover, the HBCD neonatal protocol prioritized T2-weighted acquisition, with the T1-weighted sequence collected last, T1-weighted volumetric data were available for <50% of the sample and were therefore not included in analyses ^26^. Imaging data underwent centralized quality control and preprocessing by the HBCD Data Coordinating Center. Native-space T2-weighted brain segmentations and masks were generated using BIBSNet, a deep learning model optimized for infant MRI brain tissue segmentation ^46^. Anatomical ROIs for T2-weighted images were derived from BIBSNet segmentations. dMRI data were processed with QSIRecon ^47^.

Preprocessed diffusion data were reconstructed using the QSIRecon-DIPYDKI pipeline to estimate diffusion tensor metrics, including fractional anisotropy (FA) and mean (MD), radial (RD), and axial (AD) diffusivity ^48,49^. White matter diffusion metrics were summarized within canonical white matter bundles defined using DSI Studio ^50^. Neuroimaging measures were harmonized across acquisition sites using ComBat, an empirical Bayes method that removes systematic batch effects while preserving biological variability, using the sva R package ^51^. Site (scanner) was modeled as the batch variable, and biological covariates (cannabis exposure, sex, and age [and total brain volume, where applicable]) were included in the design matrix to ensure that variance associated with these factors was retained during harmonization. Harmonized values were used in subsequent analyses.

### Region Selection and Data Preparation

Analyses targeted regions implicated in PCE and early neurodevelopment (eFigures 3-4 in the Supplement) ^52,53^, including meso- and fronto-limbic structures (amygdala, hippocampus, nucleus accumbens), basal ganglia and thalamus, cerebral gray and white matter and association regions (corpus callosum, superior and inferior longitudinal fasciculi, cingulum bundles, uncinate fasciculi), and cerebellar structures (gray and white matter, peduncles, and vermis). Neuroimaging metrics were winsorized to three standard deviations above or below the mean to account for extreme values.

### Statistical Analysis

Analyses were conducted in RStudio ^54^. Group differences in sociodemographics were assessed using independent samples t-tests and Pearson correlations. Covariate-balanced propensity scores (CBPS) were estimated using the CBPS package to balance sociodemographic variables, maternal mental health, and co-occurring substance exposures (eFigure 5 in the Supplement) ^55–57^. Linear regression models evaluated associations between PCE and neonatal MRI across (1) PCE presence in the full sample; (2) timing of PCE (first trimester versus sustained) in the full sample; and (3) frequency of PCE within the PCE subgroup. For (2), first trimester and sustained PCE were mutually exclusive and were included in the same model. For (3), sociodemographic covariates which were significantly correlated with PCE frequency were balanced by estimating a generalized propensity score for PCE frequency (eFigure 6 in the Supplement) ^55^, and PCE frequency was log-transformed to address skewness, consistent with a Box-Cox transformation analysis ^58,59^. Models were adjusted for infant sex and age at scan (corrected to 40 weeks’ gestation), with propensity scores applied as weights. As the final analytic sample contained no multiple-gestation pairs or siblings, observations were statistically independent. Volumetric models were adjusted for total brain volume. Effect sizes (partial eta squared - η^2^p), regression coefficients, and false discovery rate-corrected P-values (*P_corr_*) were reported. Given the sample size and 11-12 covariates per model, the study had greater than 80% power to detect small effect sizes (Cohen f^2^ ≈ 0.01; η^2^p ≈ 0.01) at a 2-sided α = .05.

Secondary analyses evaluated prenatal nicotine exposure (PNE) as an independent predictor alongside its interaction with PCE. Because nicotine was transitioned to an explicit predictor, we re-estimated the CBPS excluding nicotine from the balancing covariate pool. We then re-analyzed linear regressions from analytic frameworks (1)–(3) using re-estimated CBPS weights while maintaining model adjustments (infant sex, age at scan, and total brain volume for volumetric models). PNE and its interaction term with PCE were included as additional regressors; specifically, we evaluated the interaction between PNE and PCE presence in framework (1), PCE timing in framework (2), and PCE frequency within the PCE subgroup in framework (3).

## Results

### Sample Characteristics

Compared with non-exposed dyads, mothers reporting PCE had lower household income and educational attainment, were younger, and reported higher prenatal nicotine, alcohol, and opioid exposure (all *p* < .05; Table 1). Subclinical or clinical depressive symptoms (EPDS ≥10) were reported by 318 (19.5%); mild or greater anxiety symptoms (APA Cross-Cutting Anxiety, Level 1 > 1) were reported by 381 (23%).

**Table 1.** Sample Demographic Characteristics of Mother-Child Dyads with and Without PCE.

| Characteristic | No PCE<br>N = 1,408<sup>1</sup> | PCE<br>N = 222<sup>1</sup> | P-value<sup>2</sup> |
| --- | --- | --- | --- |
| Median Household Income Bracket (USD) | \$100,000 - \$199,999 | \$10,000 - \$24,999 | < .0001 |
| Educational Attainment (Years) | 18.0 (2.7) | 15.0 (2.4) | < .0001 |
| Birth Mother Age (Years) | 33.0 (5.2) | 29.6 (5.8) | < .0001 |
| % with Nicotine Exposure | 6.3% | 45% | < .0001 |
| % with Alcohol Exposure | 4.7% | 15% | < .0001 |
| % with Opioid Exposure | 3.1% | 12% | < .0001 |
| % with First Trimester Cannabis Exposure | -- | 42% | < .0001 |
| % with Sustained Cannabis Exposure | -- | 56% | < .0001 |
| Cannabis Exposure Frequency<br>(# Reported Uses over 7-week TLFB) | 1.7 (9.5) | 100.9 (105.9) | < .0001 |
| Edinburgh Postnatal Depression Score | 4.0 (4.6) | 8.0 (6.1) | < .0001 |
| APA Cross-Cutting Anxiety, Level 1 | 1.0 (1.1) | 2.0 (1.4) | < .0001 |
| Infant Age at MRI<br>(Prematurity - Adjusted, Weeks) | 3.0 (1.8) | 3.0 (2.1) | >.9 |
| Infant Gestational Age at Birth (Weeks) | 38.3 (1.8) | 37.9 (1.7) | .4 |
| Infant Sex | 613 F / 718 M<br>(77 unknown) | 106 F / 103 M<br>(13 unknown) | .3 |

<sup>1</sup> Mean (SD) <sup>2</sup> Welch Two Sample t-test, Pearson's Chi-squared test

### PCE and Neonatal Brain Structure

In full-sample analyses, PCE presence was associated with volumetric and diffusion differences in several regions (Table 2, Figure 1, *p_corr_* < .05). Specifically, PCE was associated with smaller volumes in the left and right thalamus and cerebellar vermis and larger volumes in the left hippocampus, right pallidum, left caudate, and left cerebellar cortex, as well as higher RD in the right uncinate fasciculus and superior cerebellar peduncle. While effect sizes were small across analyses, the largest effect was in the left thalamus (η^2^p > .03).

**Figure 1.**
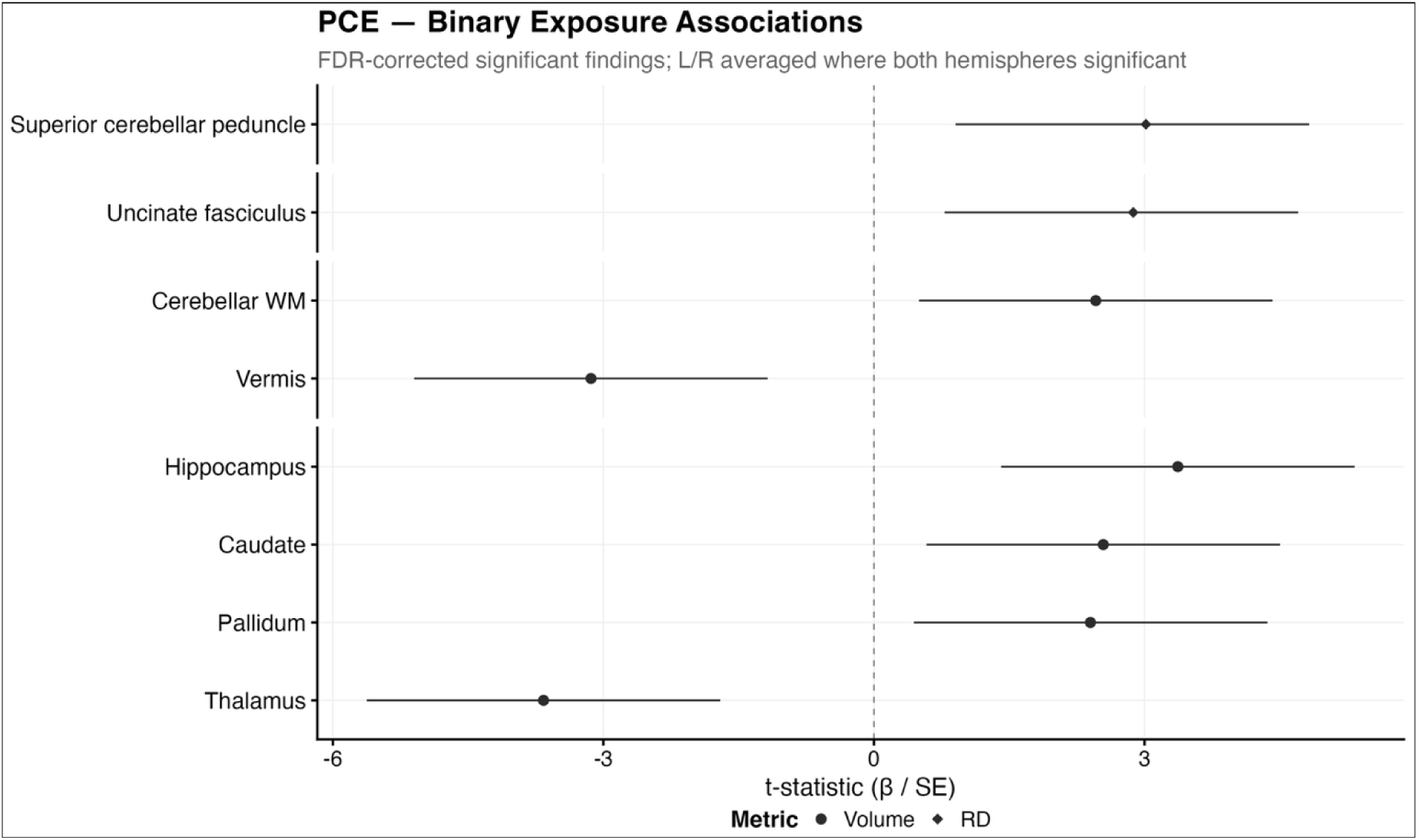
Associations Between PCE and Neonatal Brain Structure. Forest plot displaying standardized effect estimates (β/SE) for associations between PCE and white matter diffusion metrics (bottom panels) and regional brain volumes (top panels. Points represent effect estimates and horizontal lines indicate 95% confidence intervals. Shape denotes imaging metric (Volume, volumetric measure; RD, radial diffusivity). Hemisphere-specific labels were removed for visualization, and estimates were averaged when significant associations were present in both hemispheres. Positive values indicate greater regional volume or diffusivity associated with PCE, whereas negative values indicate lower values associated with PCE.

**Table 2.** Associations Between Presence of PCE and Regional Brain Measures. Parameter estimates (β), partial eta squared (η^2^p), and false discovery rate-correctedp values (P_corr_) for all statistically significant associations between PCE status and regional T2-weighted brain volumes and DTI metrics are presented. Only associations surviving FDR correction (P_corr_ < .05) are shown.

| Parameter | Region | Parameter<br>Estimate<br>($\beta$) | 95% Confidence<br>Interval | Effect<br>Size<br>($\eta^2p$) | P | $P_{corr}$ |
| --- | --- | --- | --- | --- | --- | --- |
| RD<br>(mm<sup>2</sup>/s) | R. Uncinate Fasciculus | .000011 | [.000003, .000018] | .003 | .004 | .028 |
|  | Superior Cerebellar<br>Peduncle | .000010 | [.000003, .000016] | .005 | .003 | .028 |
| Volume<br>(mm<sup>3</sup>) | R. Pallidum | 6.35 | [1.17, 11.54] | <.001 | .016 | .040 |
|  | L. Caudate | 25.01 | [5.73, 44.29] | <.001 | .011 | .038 |
|  | R. Thalamus Proper | -49.65 | [-83.07, -16.24] | .023 | .004 | .015 |
|  | L. Thalamus Proper | -71.89 | [-103.50, -40.29] | .038 | < .001 | <.001 |
|  | L. Hippocampus | 23.00 | [9.62, 36.38] | .002 | < .001 | .007 |
|  | L. Cerebellar White Matter | 236.23 | [48.00, 424.47] | <.001 | .014 | .039 |
|  | Cerebellar Vermis | -35.47 | [-57.63, -13.32] | .028 | .002 | .010 |

### Gestational Timing of PCE

Timing-specific analyses compared infants with first trimester PCE to those with sustained PCE (Tables 3-4, Figure 2). These groups showed differing volumetric associations across overlapping regions. First trimester exposure was associated with smaller left thalamus volume, whereas sustained PCE was associated with larger right and left cerebellar cortex, left caudate, right and left hippocampus volumes, and smaller right and left thalamus, cerebellar vermis, and right nucleus accumbens volumes. Effect sizes were small across analyses but were largest (η^2^p > .01) in regions where exposure was associated with smaller volumes.

**Table 3.** Associations Between First Trimester PCE and Regional Brain Measures. Parameter estimates (β). partial eta squared (pp). andfalse discovery rate-correctedp values (P_corr_) for all statistically significant associations between First Trimester PCE and regional T2-weighted brain volumes and DTI metrics are presented. Only associations surviving FDR correction (P_corr_ < .05) are shown.

| Parameter | Region | Parameter<br>Estimate ($\beta$) | 95% Confidence<br>Interval | Effect<br>Size<br>($\eta^2p$) | P | $P_{corr}$ |
| --- | --- | --- | --- | --- | --- | --- |
| MD<br>(mm<sup>2</sup>/s) | Superior Cerebellar<br>Peduncle | .000016 | [.000007, .000026] | .007 | <.001 | .013 |
| RD<br>(mm<sup>2</sup>/s) | Middle Cerebellar<br>Peduncle | .000016 | [.000006, .000027] | .004 | .002 | .019 |
|  | Superior Cerebellar<br>Peduncle | .000018 | [.000010, .000026] | .012 | <.001 | <.001 |
| Volume<br>(mm<sup>3</sup>) | L. Thalamus Proper | -76.72 | [-117.62, -35.83] | .017 | <.001 | .004 |

**Table 4.**
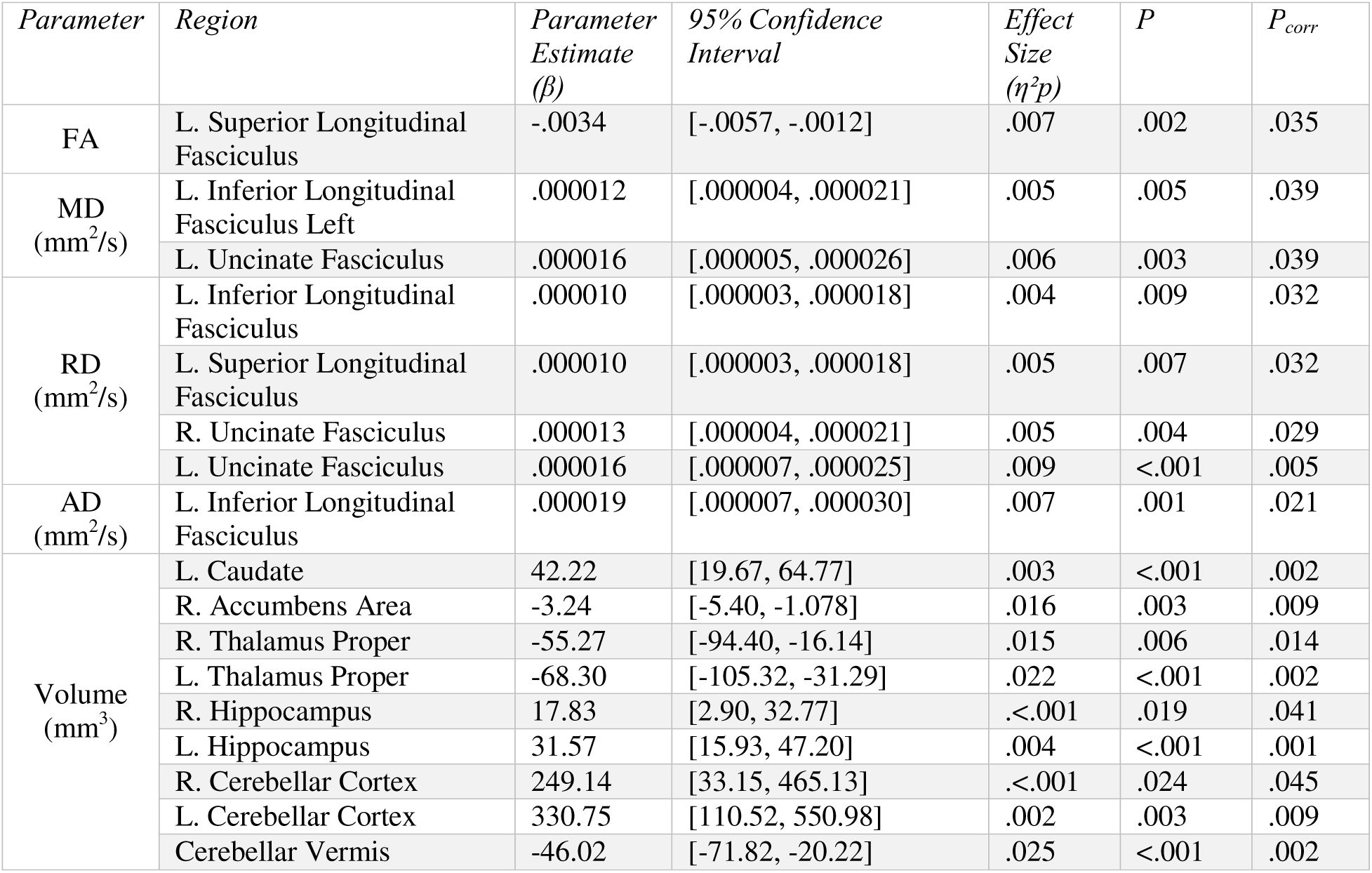
Associations Between Sustained PCE and Regional Brain Measures. Parameter estimates (β). partial eta squared (pp). andfalse discovery rate-correctedp values (P_corr_) for all statistically significant associations between Sustained PCE and regional T2-weighted brain volumes and DTI metrics are presented. Only associations surviving FDR correction (P_corr_ < .05) are shown.

**Figure 2.**
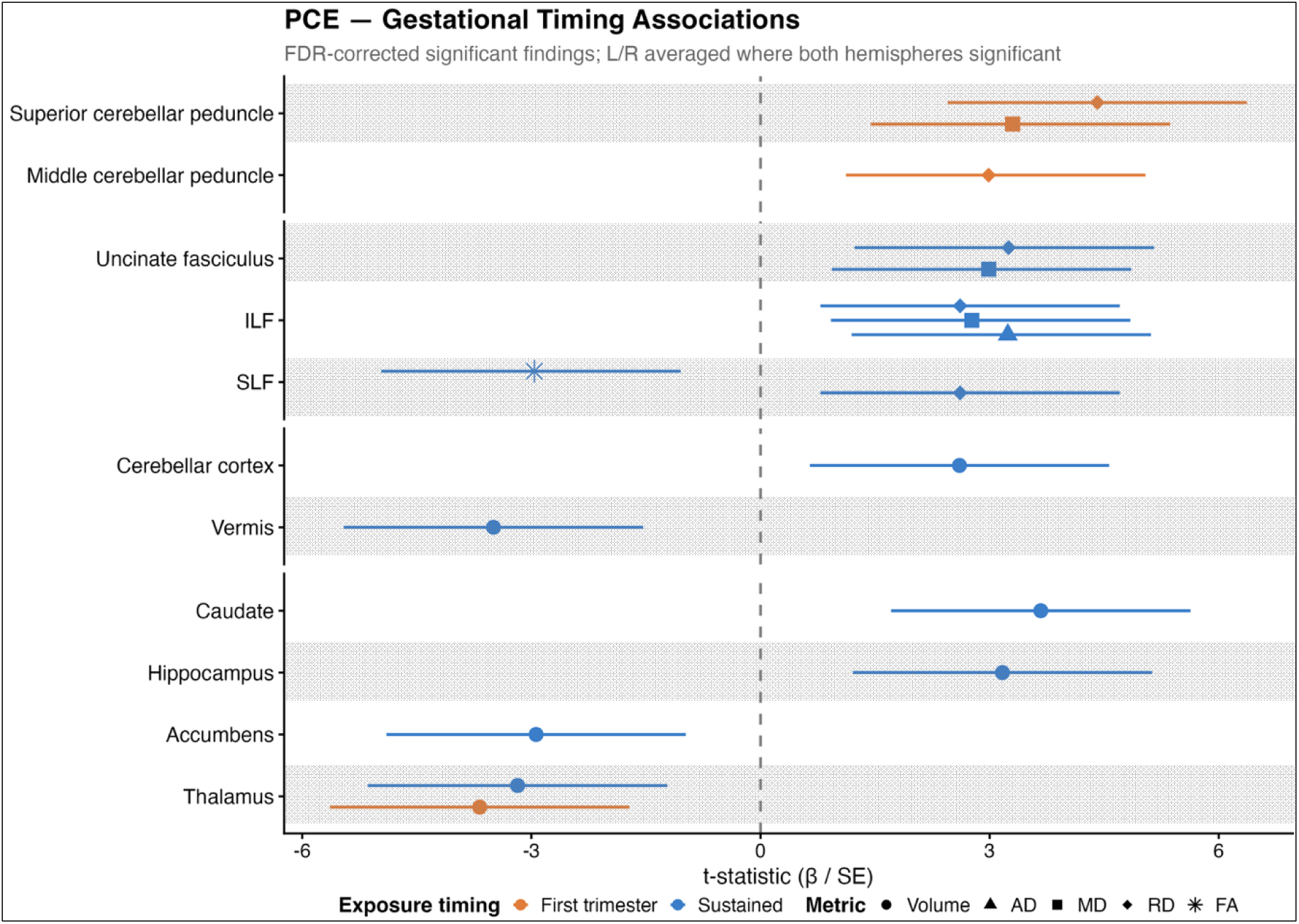
Associations Between Gestational Timing of PCE and Neonatal Brain Structure. Forest plot displaying standardized effect estimates (β/SE) for associations between first trimester (orange) and sustained (blue) PCE and white matter diffusion metrics (bottom panels) and regional brain volumes (top panels. Points represent effect estimates and horizontal lines indicate 95% confidence intervals. Shape denotes imaging metric (Volume, volumetric measure; MD, mean diffusivity; RD, radial diffusivity; AD, axial diffusivity; FA, fractional anisotropy). Hemisphere-specific labels were removed for visualization, and estimates were averaged when significant associations were present in both hemispheres. Positive values indicate greater regional volume or diffusivity associated with PCE, whereas negative values indicate lower values associated with PCE.

Diffusion MRI analyses revealed timing-specific microstructural associations. First trimester exposure was associated with higher diffusivity values in the superior (MD and RD) and middle (RD) cerebellar peduncles. Sustained exposure was associated with lower FA in the left superior longitudinal fasciculus and higher diffusivity values in the left superior longitudinal fasciculus (RD), right (RD) and left (MD and RD) uncinate fasciculi and left inferior longitudinal fasciculus (MD, RD, and AD). Effect sizes were small (η^2^p < .02) across significant regions.

### Frequency-Dependent Associations Within the PCE Subgroup

Within the PCE-exposed subgroup, greater frequency of exposure was associated with larger right and left amygdala, left hippocampus, and left cerebral white matter volume (small effect sizes) and higher diffusivity values in the right (RD) and left (MD and RD) uncinate fasciculi, right (MD and RD) cingulum bundle, and right and left superior longitudinal fasciculi (RD) with medium-to-large effect sizes (η^2^p > .1), Table 5, Figure 3).

**Table 5.** Associations Between PCE Frequency and Regional Brain Measures. Parameter estimates (β). partial eta squared (pp). andfalse discovery rate-correctedp values (P_corr_) for all statistically significant associations between PCE frequency and regional T2-weighted brain volumes and DTI metrics are presented. Only associations surviving FDR correction (P_corr_ < .05) are shown.

| *Parameter* | *Region* | *Parameter Estimate ($\beta$)* | *95% Confidence Interval* | *Effect Size ($\eta^2p$)* | *P* | *P<sub>corr.</sub>* |
| --- | --- | --- | --- | --- | --- | --- |
| MD (mm<sup>2</sup>/s) | R. Cingulum | .000010 | [.000003, .000016] | .13 | .004 | .036 |
|  | L. Uncinate Fasciculus | .000014 | [.000004, .000024] | .10 | .005 | .036 |
| RD (mm<sup>2</sup>/s) | R. Cingulum | .000009 | [.000003, .000015] | .13 | .004 | .027 |
|  | R. Superior Longitudinal Fasciculus | .000009 | [.000003, .000016] | .13 | .007 | .027 |
|  | L. Superior Longitudinal Fasciculus | .000009 | [.000002, .000016] | .11 | .016 | .047 |
|  | R. Uncinate Fasciculus | .000012 | [.000004, .000021] | .12 | .005 | .027 |
|  | L. Uncinate Fasciculus | .000014 | [.000005, .000022] | .12 | .002 | .027 |
| Volume (mm<sup>3</sup>) | R. Amygdala | 8.10 | [2.28, 13.92] | .008 | .007 | .028 |
|  | L. Amygdala | 7.96 | [2.63, 13.29] | .007 | .004 | .020 |
|  | L. Cerebral White Matter | 720.87 | [346.76, 1094.97] | .004 | <.001 | .003 |
|  | L. Hippocampus | 29.31 | [10.81, 47.82] | .013 | .002 | .017 |

**Figure 3.**
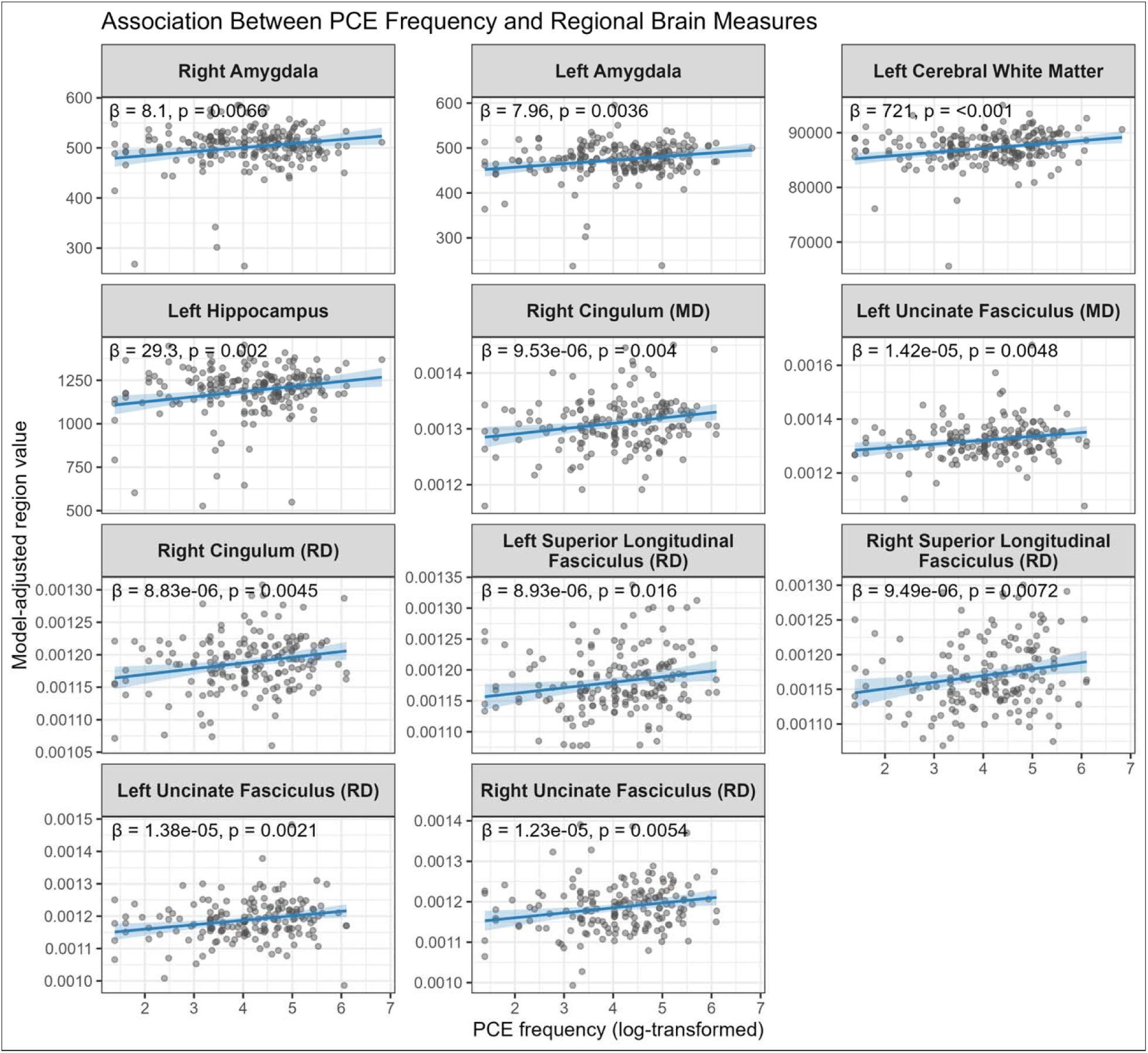
Associations Between PCE Frequency and Regional Brain Measures. Significant associations between frequency of PCE and T2-weighted volumes and DTI Metrics. Lines show model-adjusted predictions (±95% CI) from weighted linear regressions; points are observed values plus residuals. Volumetric models adjusted for total brain volume, sex, and age; MD (mean diffusivity) and RD (radial diffusivity) models adjusted for sex and age. β and p-values are for the PCE frequency coefficient.

Across analyses, infant age (corrected to a 40-week gestation) and sex were included as covariates. Infant sex did not moderate associations between PCE and brain measures, and nonsignificant interaction terms were removed to improve model fit based on AIC.

### Interactions between PCE and PNE

We next explicitly included PNE and in our analytic framework, as 45% of our PCE subsample reported PNE (Table 1). When PNE and the PCE×PNE interaction were included in framework (1), previously significant volumetric associations between PCE presence and neonatal brain volumes were attenuated to non-significance except in the cerebral white matter. PNE emerged as an independent predictor of volume across several regions, with significant PCE×PNE interactions observed in the hippocampus, pallidum, caudate, cerebral white matter, thalamus, and cerebellar cortex (all small effect sizes, η^2^p < .01) In contrast, white matter diffusion microstructural associations with PCE remained significant in the inferior longitudinal fasciculi, uncinate fasciculi, cerebellar white matter, and superior cerebellar peduncle (with a significant PCExPNE interaction). PNE was not independently associated with diffusion microstructure (eFigure 7 in the Supplement).

In framework (2), timing-specific associations remained significant across both volumetric and diffusion metrics after inclusion of PNE. First trimester PCE was associated with smaller volumes and higher diffusivity and lower FA values in the cerebellum and inferior longitudinal fasciculi. Sustained PCE remained associated with volumes in select regions, along with previously reported diffusion associations in white matter association regions. Significant PCE-PNE interactions emerged across both first trimester and sustained use (eFigure 8-9 in the Supplement). In framework (3), frequency-dependent associations with PCE remained significant for diffusion metrics: greater frequency of use was associated with higher diffusivity values in the inferior and superior longitudinal fasciculi, uncinate fasciculi, cingulum bundles, corpus callosum, and cerebellar peduncles and white matter. However, volumetric associations within the PCE subgroup were attenuated to non-significance with the inclusion of PNE (eFigure 10 in the Supplement).

## Discussion

Increasing rates of PCE have raised public health concerns ^60^, yet associations with brain development remain incompletely characterized ^61,62^. We found that the most robust and exposure-specific associations of PCE were in white matter microstructure, whereas subcortical volumetric differences were attenuated after accounting for co-occurring nicotine exposure.

Across PCE presence, timing, frequency, and PNE-adjusted models, PCE was associated with higher diffusivity in white matter regions. Dose-response relationships linked higher exposure frequency to higher diffusivity values in the superior longitudinal fasciculi, uncinate fasciculi, and cingulum bundles, supporting a potentially exposure-specific association. Timing analyses suggested additional specificity: first-trimester exposure was associated with cerebellar white matter differences, whereas sustained exposure was linked to later developing long-range association pathways, patterns that persisted after accounting for PNE ^63,64^. These findings are in line with region-specific trajectories of myelination in infants ^65^ and suggest that exposure timing may capture distinct neurodevelopmental processes rather than simply cumulative dose. Associations observed for first-trimester but not sustained exposure could also relate to effects of abrupt cessation of use, as suggested in prior work ^66^.

Given the prolonged postnatal maturation of white matter, early diffusion differences may precede later alterations in structural connectivity or function ^67–69^. Higher diffusivity values may reflect reduced axonal coherence, delayed pre-myelination, or other alterations in early white matter organization, although diffusion tensor imaging cannot distinguish among these underlying cellular processes ^70^. These interpretations are biologically plausible given evidence that endocannabinoid signaling regulates oligodendrocyte maturation, axonal guidance, and other processes critical for white matter development ^10,22^. The sparse associations observed with fractional anisotropy may reflect the developmental stage of the sample as white matter is largely unmyelinated in neonates, and fractional anisotropy is sensitive to myelination ^65^.

Many of the regions identified here exhibit high fetal cannabinoid receptor expression ^71^, suggesting potential vulnerability to PCE. In animal models, PCE has been associated with disrupted cerebellar development and altered white matter maturation ^72^. PCE has similarly been linked to altered structural and functional connectivity during infancy in the cerebellum and frontal white matter ^18,19^ and differences in fronto-limbic white matter microstructure in older children ^22^. These findings suggest that the neonatal white matter alterations observed here may represent early manifestations of disrupted endocannabinoid-mediated neurodevelopment ^62^.

In contrast to the consistent white matter findings, volumetric associations showed greater sensitivity to co-occurring nicotine exposure. Prior to accounting for PNE, PCE was associated with differences in subcortical and cerebellar brain volumes, but volumetric associations were largely attenuated after adjustment for PNE, and significant PCE×PNE interactions emerged across multiple regions, suggesting joint or synergistic effects. In contrast, some timing-specific volumetric associations persisted after accounting for PNE. Thus, volumetric associations in overall exposure analyses may be influenced by co-occurring nicotine exposure, whereas some timing-specific associations appear less sensitive to nicotine adjustment. Overlapping subcortical targets of cannabinoid and nicotinic signaling further support the possibility of shared or interacting neurobiological pathways ^73^. Differences in cannabinoid receptor signaling across neuronal populations and glial cells may also contribute to differential interactions between PCE and PNE and associations with subcortical volumetric versus white matter trats, given the roles of endocannabinoid signaling in synaptic development and oligodendrocyte maturation, although these mechanisms cannot be distinguished with MRI alone ^10^.

The regions implicated in PCE, including the cingulum bundles, uncinate fasciculi, inferior longitudinal fasciculi, superior longitudinal fasciculi, and cerebellar white matter, support a broad range of cognitive, emotional, language, attentional, and motor functions. The cingulum bundles and uncinate fasciculi have been implicated in executive function, emotional regulation, and limbic–prefrontal integration ^74–77^, while the inferior and superior longitudinal fasciculi contribute to associative processing, language, and attention ^78,79^. Cerebellar white matter also plays an important role in motor coordination and higher-order cognitive processes ^80–83^. Future longitudinal studies should determine whether the white matter differences observed here are associated with later neurobehavioral outcomes across functional domains. Such work would extend findings from studies of adolescents with PCE, which have reported differences in executive function, attention, emotional regulation, and reward-related behaviors ^4,84^. Alternatively, early white matter differences may attenuate or normalize over time.

Notably, we did not adjust for race or ethnicity, as they are imperfect proxies for social and structural processes, and instead adjusted for directly measured family socioeconomic characteristics and maternal mental health. This approach does not capture unmeasured structural factors, including experiences of discrimination, that may contribute to the observed associations, and residual confounding cannot be excluded. Furthermore, although we used covariate balancing to reduce differences between exposed and unexposed groups, PCE occurs within a broader constellation of biological, behavioral, genetic, and social factors that also influence fetal brain development. Co-occurring substance use ^53^, maternal mental health ^85^, socioeconomic adversity ^86,87^, and nutrition ^88,88^ have each been associated with variation in brain structure or white matter development and may contribute to the observed associations independent of, or in conjunction with, PCE. Future studies that explicitly model these co-occurring influences may contribute to our understanding of how PCE influences neurodevelopmental risk in the real-world context. Additionally, individuals who discontinue cannabis use after pregnancy recognition may differ systematically from those who continue use throughout pregnancy with respect to these and other characteristics, complicating interpretation of timing-specific findings. Thus, the observed associations may reflect the combined influence of PCE and correlated environmental and maternal factors rather than any single exposure.

### Strengths and Limitations

This study has several strengths. To our knowledge, this is the largest neuroimaging study to-date examining infant brain development in the setting of PCE. Leveraging a large, well-characterized, multisite cohort, we evaluated PCE using complementary exposure definitions, enabling assessment of both the presence and pattern of exposure. We also explicitly accounted for nicotine use (PNE) and examined nicotine-by-cannabis interactions, strengthening inference regarding exposure specificity. Finally, we compared associations across diffusion white matter microstructure and regional brain volumes, providing a more comprehensive assessment of early neurodevelopment than either modality alone.

Nonetheless, several limitations warrant consideration. Although many associations reached statistical significance and effect sizes were medium-to-large for frequency analyses, effect sizes were small for presence and timing analyses, reflecting modest variance explained. Findings should therefore be interpreted cautiously and evaluated longitudinally. Despite the use of covariate-balanced propensity scores, residual confounding remains possible ^89^. This is particularly relevant for nicotine sensitivity analyses, as residual differences between nicotine-exposed and unexposed participants may remain as groups were balanced based on PCE status. Accordingly, the nicotine main effect should not be interpreted in isolation and cannabis-nicotine interactions should be interpreted cautiously, and future studies should explicitly model the joint exposure to cannabis and nicotine.

There were also limitations to our exposure classifications. Because our definition of exposure required use in at least four weeks, and the later window (weeks 7-9) spans only three weeks, use confined to the later portion of pregnancy could not meet the threshold by construction, and no participants met criteria for late-only use. Our comparison therefore contrasts early-only use with sustained (early and later) use, and sustained use necessarily reflects a greater number of exposed weeks. Conversely, early-only reports likely include individuals who discontinued use upon learning of the pregnancy. Differences between these groups may therefore reflect frequency or duration of exposure rather than gestational timing, and we cannot distinguish these explanations. Additionally, because exposure was assessed at discrete time points, some individuals may have been misclassified with respect to timing or duration of use, potentially attenuating observed associations. Frequency analyses within exposed individuals should also be interpreted cautiously despite medium-to-large effect sizes given reduced sample size and potential heterogeneity. Future samples, including future HBCD data releases, with more granular repeated exposure measurement, information on mode of administration, and sufficient late-pregnancy limited use will be needed to disentangle timing from dose.

Additionally, although diffusion tensor imaging and volumetric MRI are sensitive markers of early neurodevelopment, they lack biological specificity, limiting conclusions regarding the cellular mechanisms (e.g., myelination, axonal organization, or glial development) underlying the observed differences ^67,90,91^. Furthermore, cortical gray and white matter analyses were limited to whole-hemisphere averages; region-specific cortical trajectories should be examined in future work. Finally, the cross-sectional design precludes inference regarding developmental trajectories, causal relationships, paternal contributions, and the functional significance of the observed brain differences.

### Future Directions

Longitudinal follow-up within HBCD will be critical to understand how brain differences relate to later developmental outcomes. It will also be important to identify periods of heightened vulnerability as well as resilience ^92,93^. Advanced imaging approaches, such as neurite orientation dispersion and density imaging ^94–96^ and quantitative relaxometry ^67,97^, may further clarify underlying neurobiology. Future work should prioritize linking structural and microstructural differences to downstream developmental outcomes to establish functional significance.

## Conclusions

Leveraging a large, well-phenotyped multisite cohort, we examined associations between PCE and early brain development. The most consistent associations emerged in white matter microstructure, which demonstrated frequency-dependent effects that were robust across analytic approaches and remained after accounting for nicotine (PNE). In contrast, subcortical volumetric differences were more sensitive to PNE, with attenuation of main effects and evidence of PCE×PNE interactions, suggesting that volumetric findings in combined exposure models may reflect polysubstance-related influences. These results indicate differential sensitivity of neonatal brain measures to prenatal cannabis and nicotine exposure. Our findings are consistent with subtle early neurodevelopmental differences whose stability and functional significance remain uncertain. Longitudinal studies are needed to determine whether these differences persist, normalize, or emerge as later cognitive and behavioral outcomes.

Clinically, these findings support developmentally informed counseling regarding cannabis use during pregnancy, with attention to timing, frequency, and the high prevalence of co-occurring nicotine exposure ^98^. However, the modest magnitude of observed effects, relative to well-established teratogens such as alcohol ^99,100^, argues against interpreting these associations as deterministic or using them in isolation for clinical decision-making. Nonjudgmental, patient-centered communication remains essential to support disclosure, reduce stigma, and facilitate shared decision-making. Overall, these results highlight subtle associations between PCE and early white matter development while underscoring the need for longitudinal research.

## Acknowledgements

We extend our sincere gratitude to the families who generously participated in this study, and to the students and research staff whose dedication and expertise made the collection of these data possible. A previous version of this manuscript was posted on medRxiv at https://doi.org/10.64898/2026.03.02.26347436. The authors used Anthropic Claude to refine, edit, and improve the clarity, grammar, and formatting of human-written text and code used in this manuscript. No AI tool was used to generate the scientific content, interpret results, or draw conclusions. All AI-generated suggestions were reviewed and evaluated by the authors, who revised the manuscript as appropriate. The authors take full responsibility for the accuracy, integrity, and content of the manuscript.

## Data Availability Statement

Data used in the preparation of this article were obtained from the <u>HEALthy Brain and Child Development (HBCD) Study</u>, held in the NIH Brain Development Cohorts Data Sharing Platform. This is a multisite, longitudinal study designed to recruit approximately 7,000 families and follow them from pregnancy to early childhood. The HBCD Study is supported by the NIH and additional federal partners under award numbers U01DA055352, U01DA055353, U01DA055366, U01DA055365, U01DA055362, U01DA055342, U01DA055360, U01DA055350, U01DA055338, U01DA055355, U01DA055363, U01DA055349, U01DA055361, U01DA055316, U01DA055344, U01DA055322, U01DA055369, U01DA055358, U01DA055371, U01DA055359, U01DA055354, U01DA055370, U01DA055347, U01DA055357, U01DA055367, U24DA055325, and U24DA055330. A full list of supporters is available at <u>Federal Partners-HBCD Study</u>. A full list of participating sites is available at <u>Study Sites-HBCD Study</u>. HBCD Study Consortium investigators designed and implemented the study and/or provided data but did not necessarily participate in the analysis or writing of this report. This manuscript reflects the views of the authors and may not reflect the opinions or views of the NIH or the HBCD Study Consortium investigators. The HBCD dataset grows and changes over time. The HBCD data used in this report came from [https://doi.org/10.82525/vnye-pe87]. DOIs can be found at [https://www.nbdc-datahub.org/hbcd-study]. Corresponding author Leela Shah had access to all data required for this investigation through an approved Data Use Certificate. R Scripts used to conduct this analysis are available at https://github.com/Developing-Brain-Imaging-Lab/Prenatal-Cannabis-Brain-Development-HBCD-Cohort.

## Supplemental Information

**eFigure 1.**
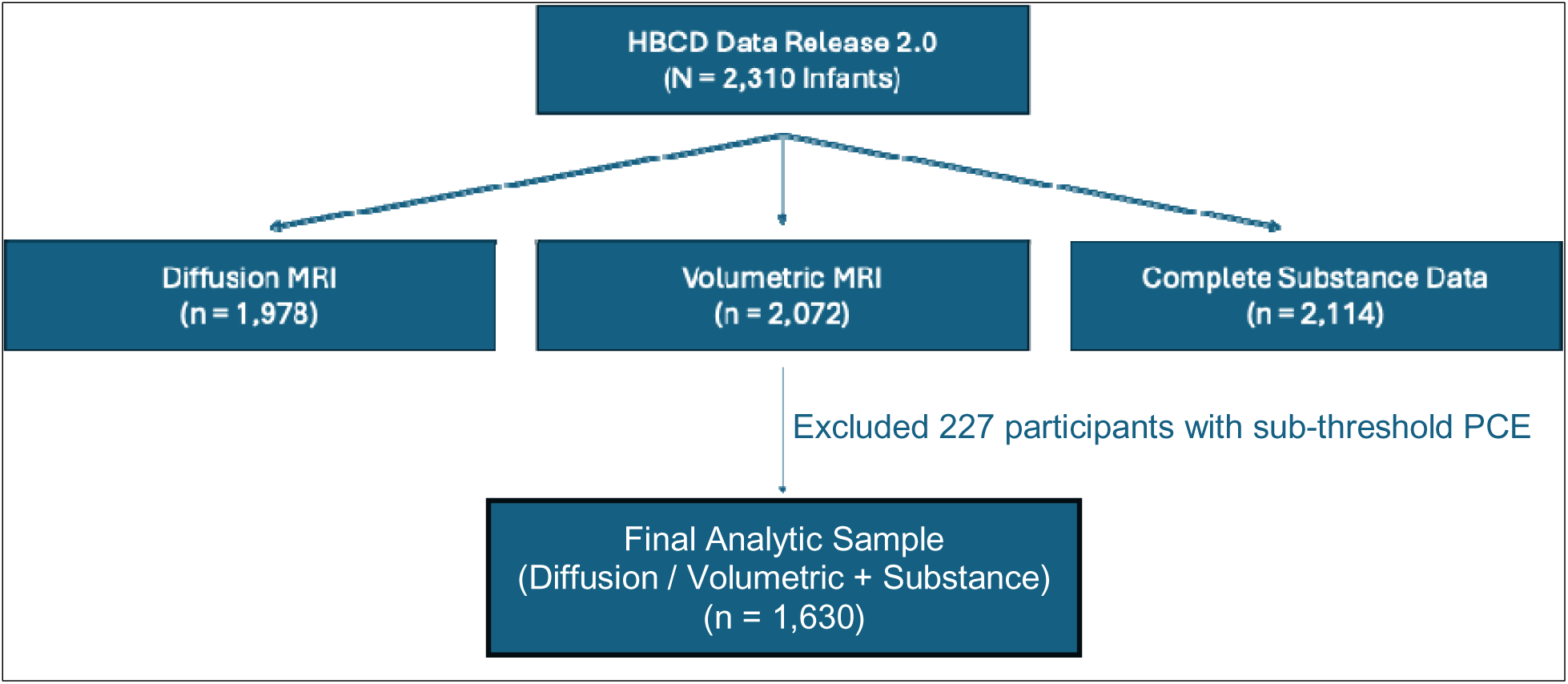
Participant Flowchart. Of 2,310 infants from the HBCD Data Release 2.0, 1,978 had diffusion MRI, 2,072 had volumetric MRI, and 2,114 had complete substance use data. 227 participants were excluded due to sub-threshold PCE. A total of 1,630 participants with complete data (either diffusion MRI or volumetric MRI + complete substance data) were included in the final analytic sample.

**eFigure 2.**
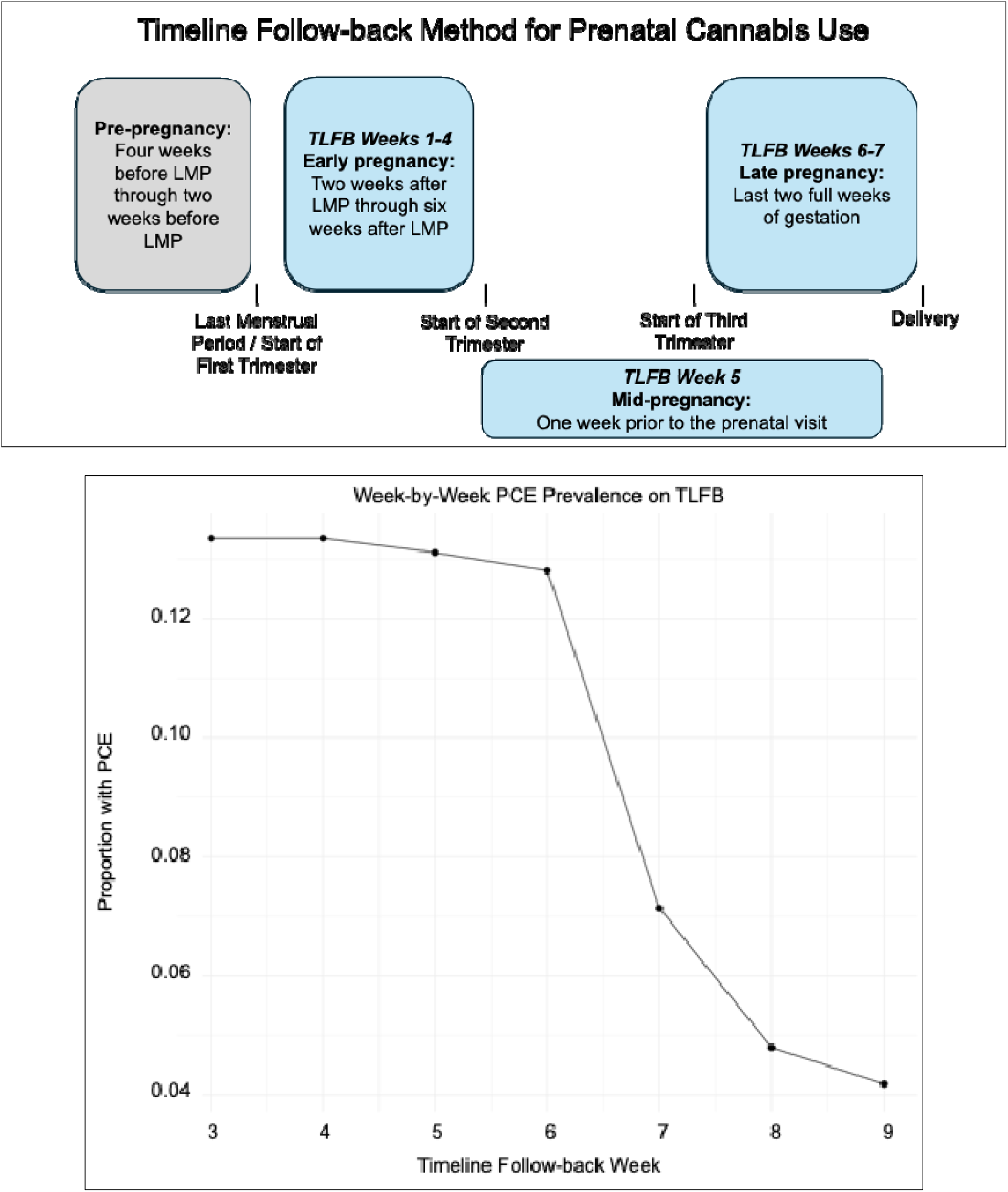
Assessment Windows for Cannabis Use Across Pregnancy. Cannabis use was retrospectively assessed using the Timeline Follow-Back (TLFB) method for the pre-pregnancy period and for early, mid, and late pregnancy based on timing relative to the last menstrual period and prenatal visits ^39^. Blue boxes indicate weeks assessed in PCE timing analyses. The graph indicates the prevalence of reported prenatal cannabis use across TLFB weeks 1-7 for participants with complete TLFB information.

**eFigure 3.**
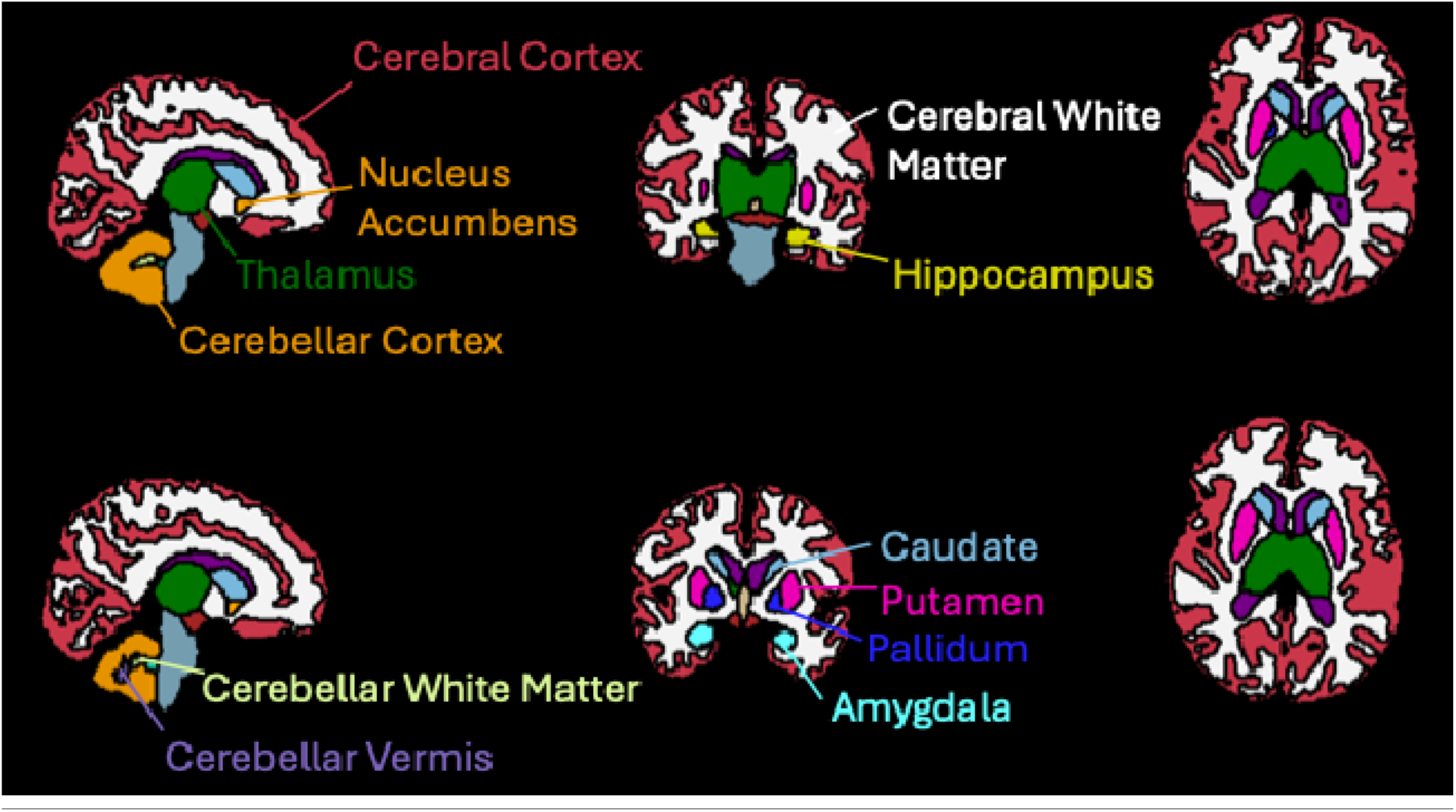
Population-Averaged Infant Volumetric Atlas. Sagittal (left), coronal (center), and axial (right) views of the atlas derived from BIBSNet segmentations co-registered to a study-specific ANTs template, visualized in FSLeyes. Regions of interest are labeled.

**eFigure 4.**
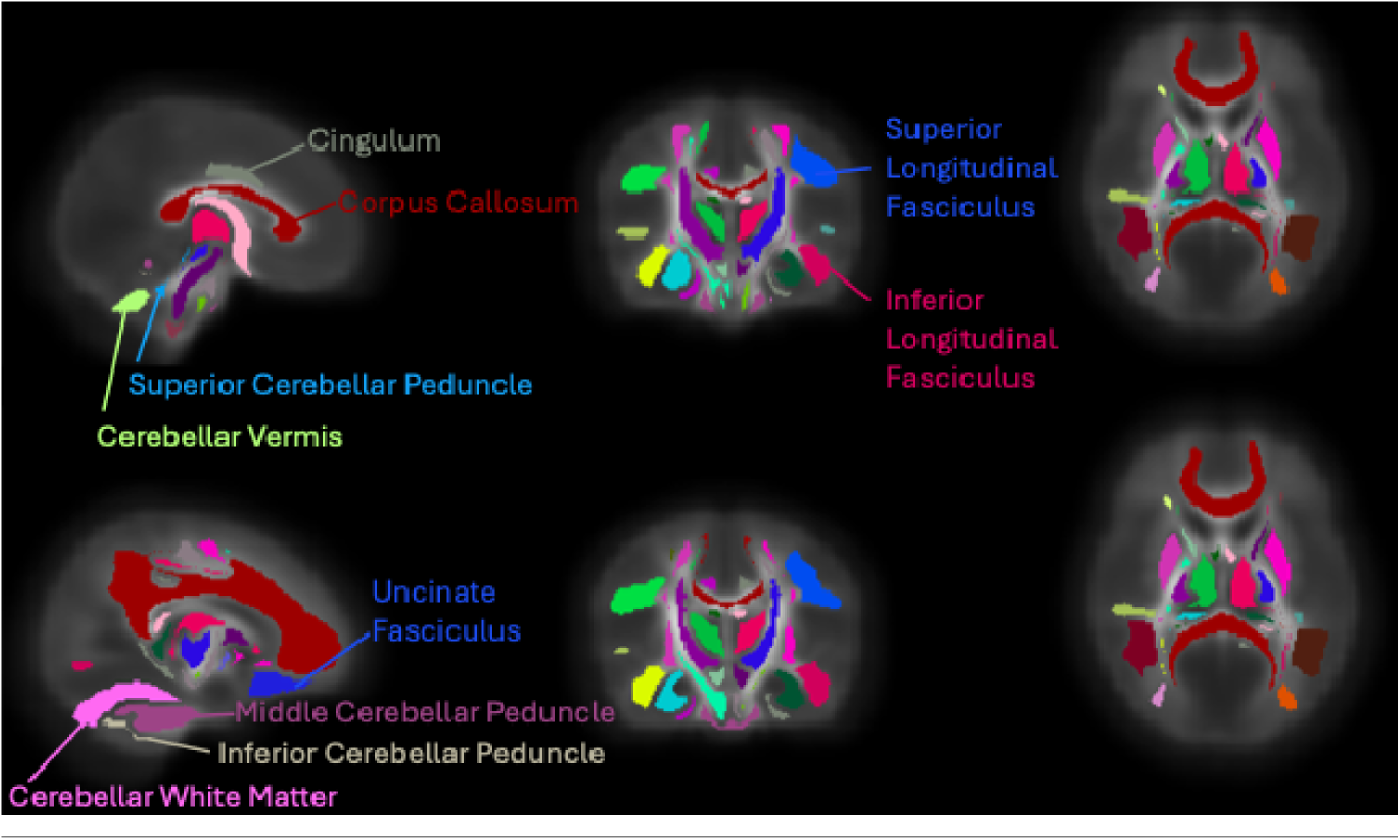
Population-Averaged Infant DKI (FA) Atlas. Sagittal (left), coronal (center), and axial (right) views of the atlas derived from dsiPrep tractography co-registered to a study-specific ANTs template, visualized in FSLeyes. Regions of interest are labeled.

**eFigure 5.**
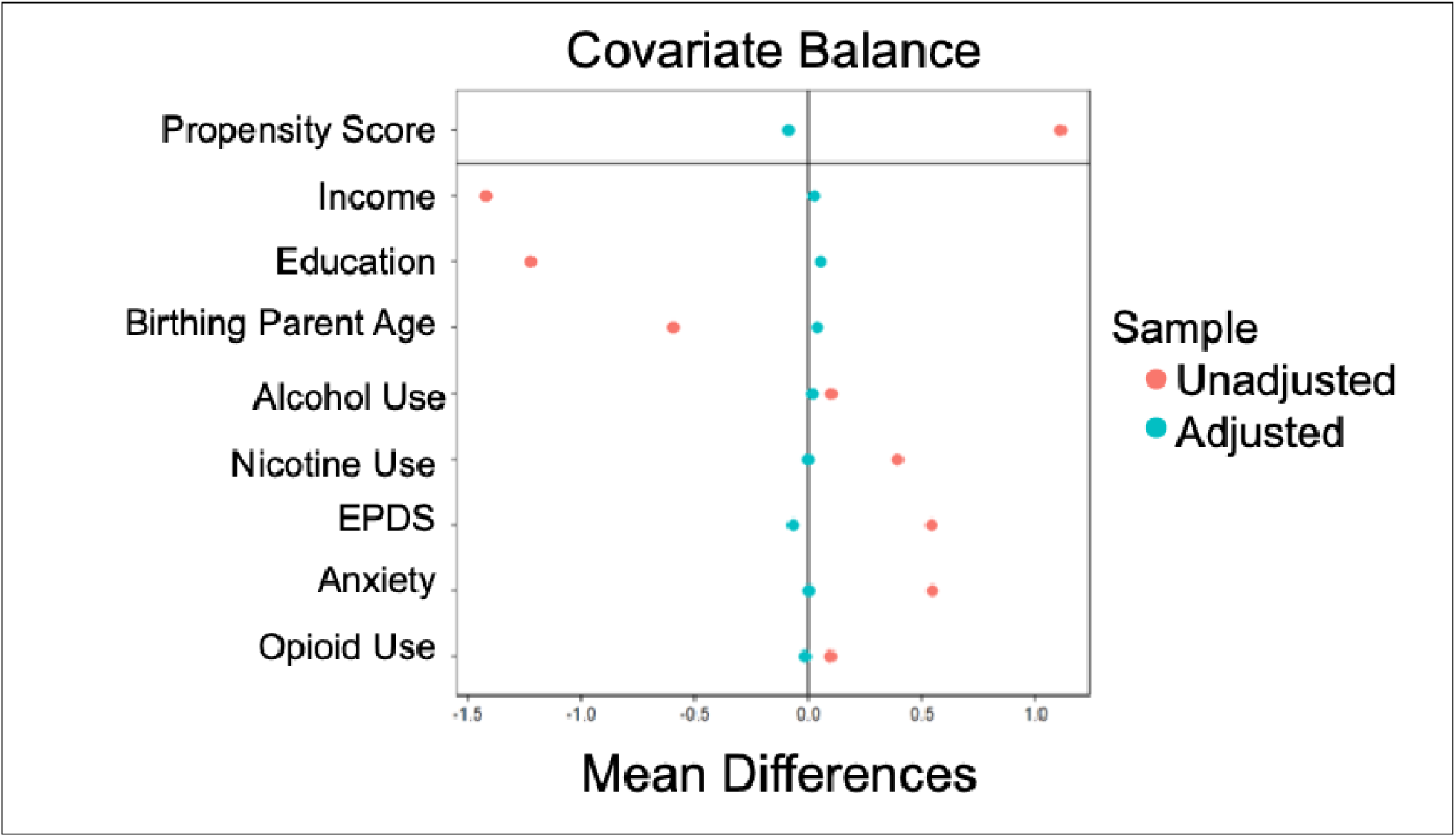
Mean Differences in Socioecological Covariates Between Parents With and Without PCE Before and After Covariate Balancing. Differences were calculated relative to the unexposed group; negative values indicate lower mean values in the PCE group. Estimates before and after covariate balancing were obtained using the covariate balancing propensity score (CBPS) method ^55^. Coral indicates values before balancing; teal, after balancing.

**eFigure 6.**
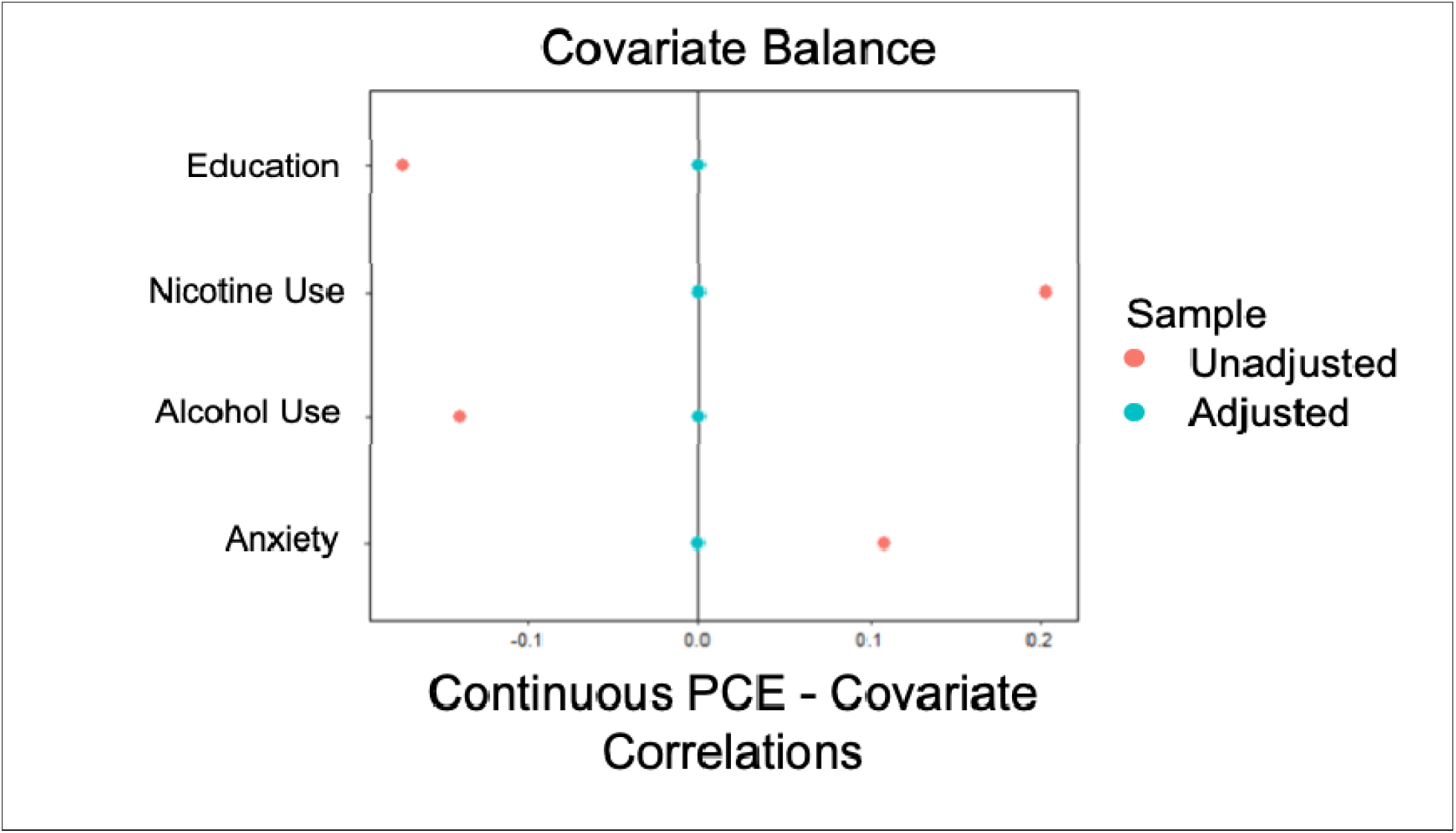
Correlations Between Socioecological Covariates and Continuous PCE Before and After Covariate Balancing. Correlations are shown for the PCE-only subgroup before and after covariate balancing using the covariate balancing propensity score (CBPS) method ^55^. Negative values indicate lower mean values for infants with higher continuous (cumulative) PCE. Coral indicates values before balancing; teal, after balancing. Only predictors that were significantly correlated (p > .05) with PCE frequency were included in the CBPS.

**eFigure 7.**
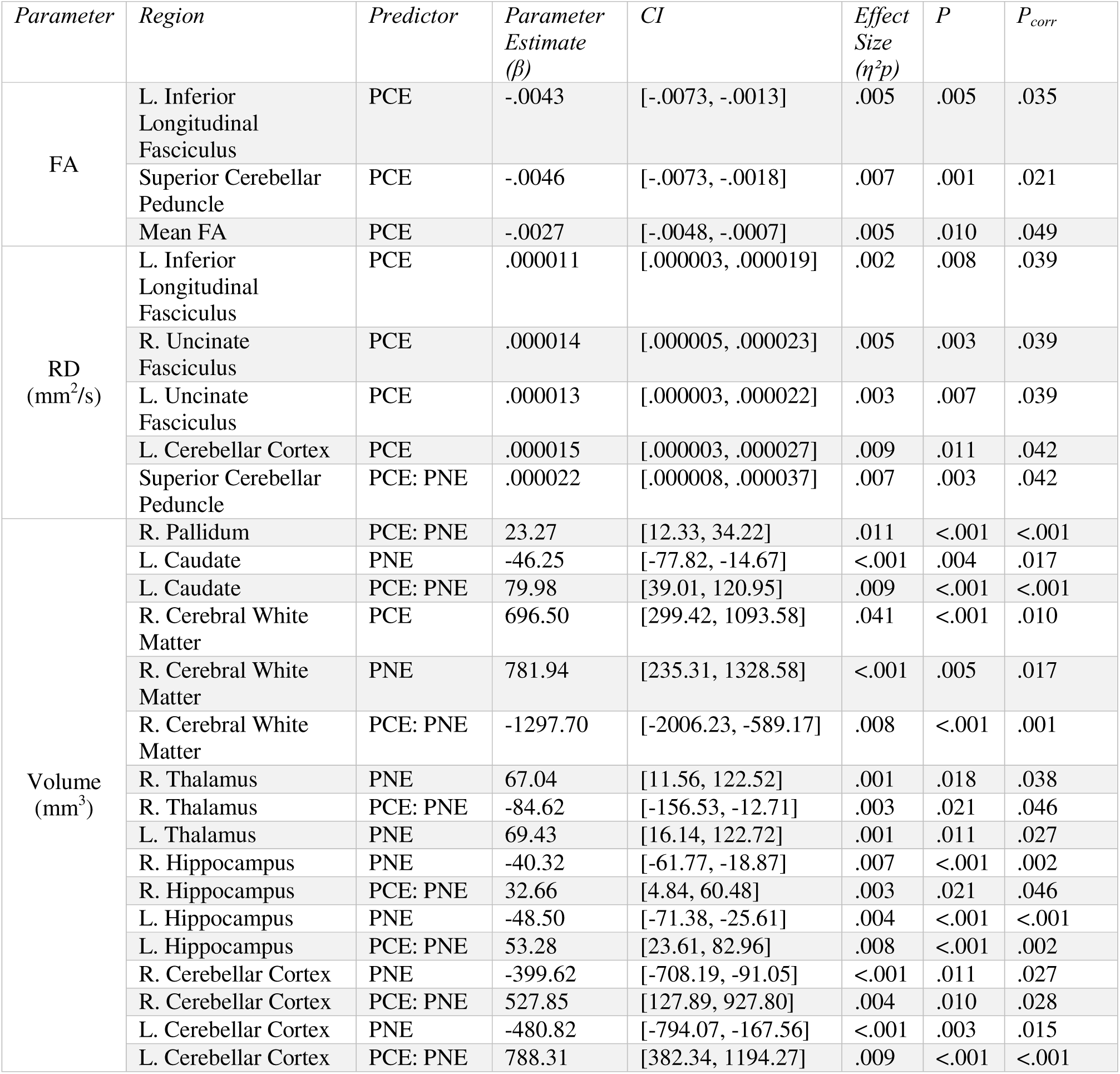
Sensitivity Analysis: Associations Between PCE Status and Regional Brain Measures, Covarying for Nicotine Use (NIC) Parameter estimates (β), partial eta squared (η^2^p), andfalse discovery rate-correctedp values (P_corr_) for all statistically significant associations between PCE status, Nicotine use (PNE), and their interaction and regional T2-weighted brain volumes and DTI metrics are presented. Only associations surviving FDR correction (P_corr_ < .05) are shown.

**eFigure 8.**
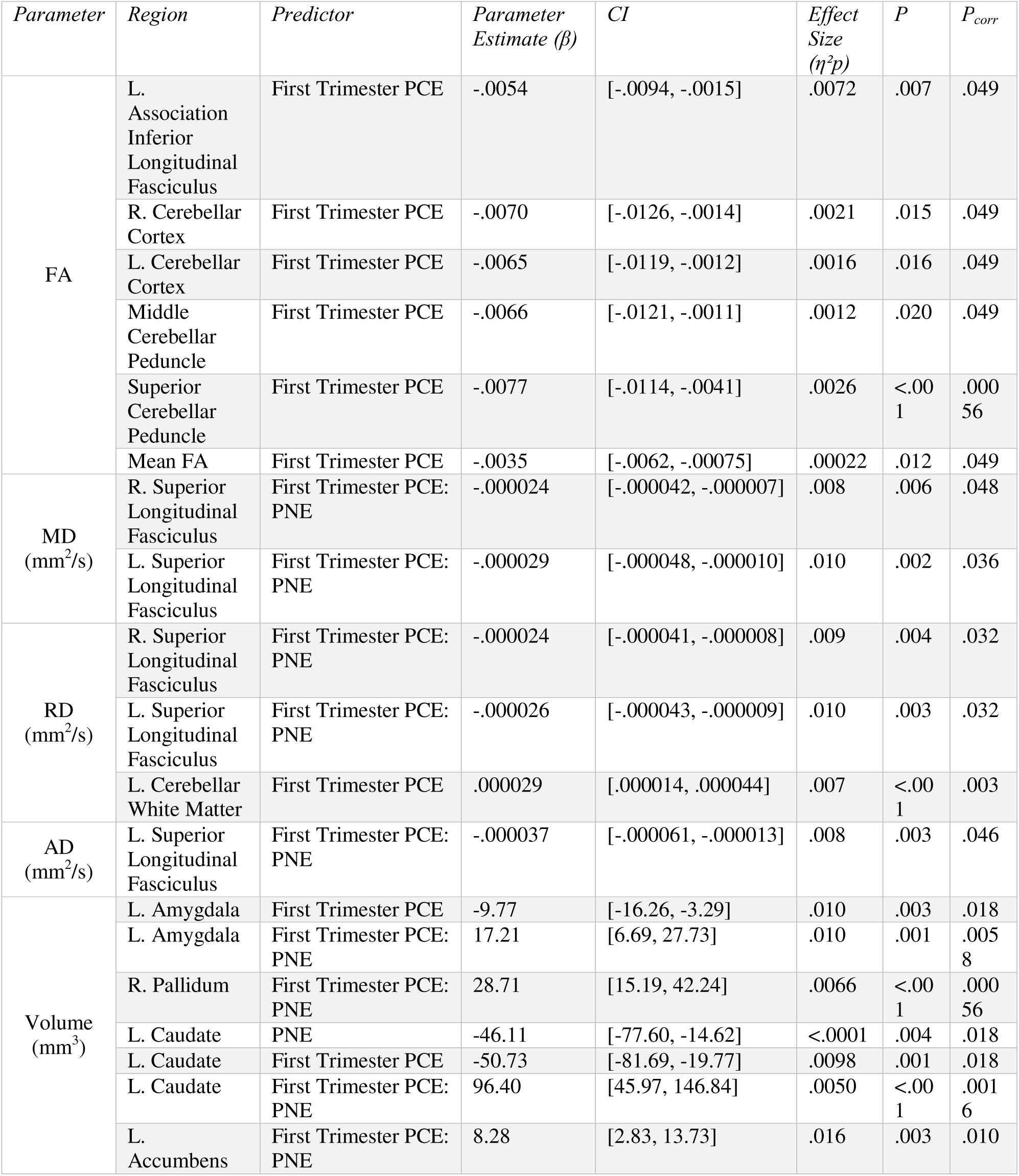
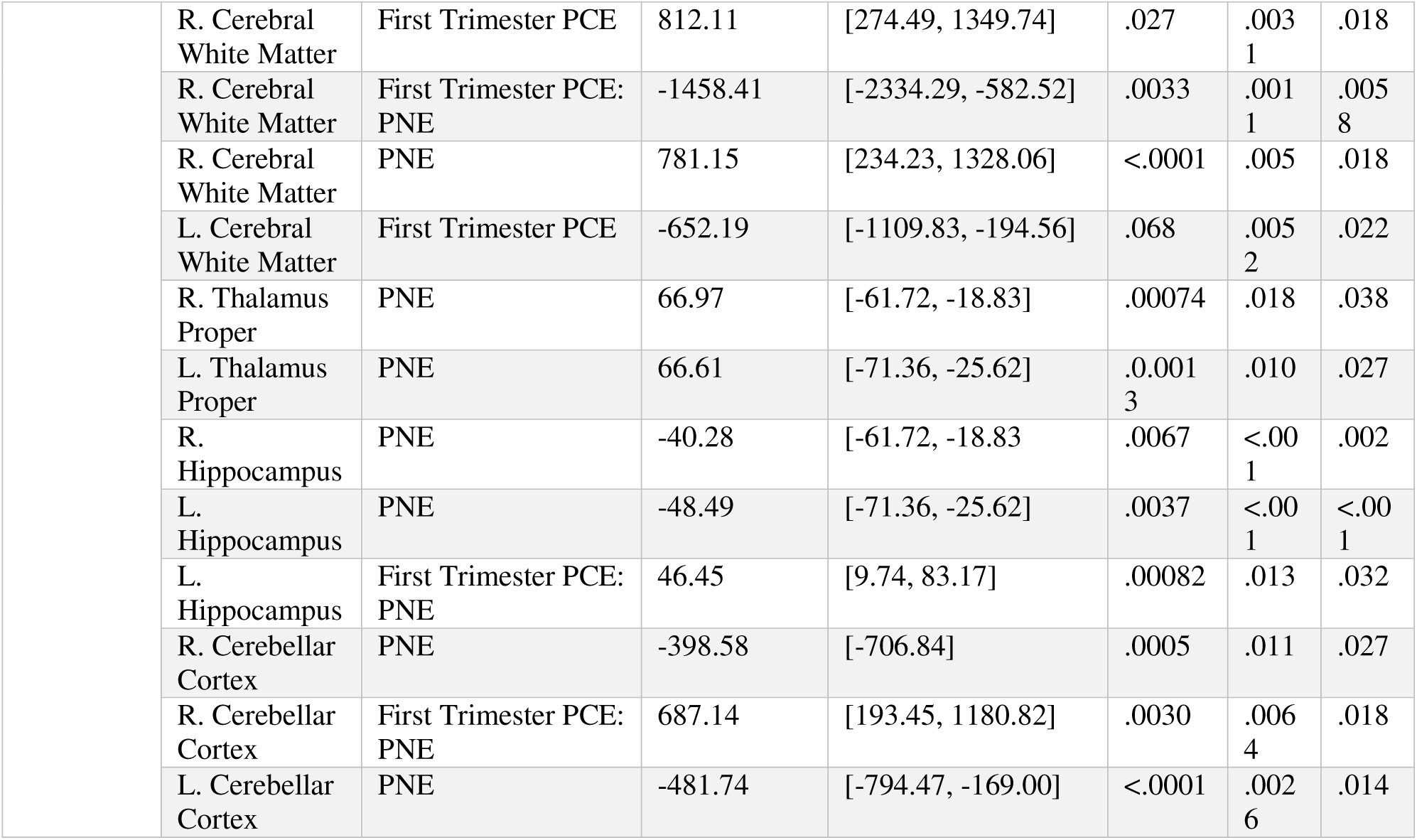
Sensitivity Analysis: Associations Between First Trimester PCE and Regional Brain Measures, Covarying for Nicotine Use (PNE) Parameter estimates (β), partial eta squared (η^2^p), andfalse discovery rate-correctedp values (P_corr_) for all statistically significant associations between First Trimester PCE, Nicotine use (PNE), and their interaction and regional T2-weighted brain volumes and DTI metrics are presented. Only associations surviving FDR correction (P_corr_ < .05) are shown.

**eFigure 9.**
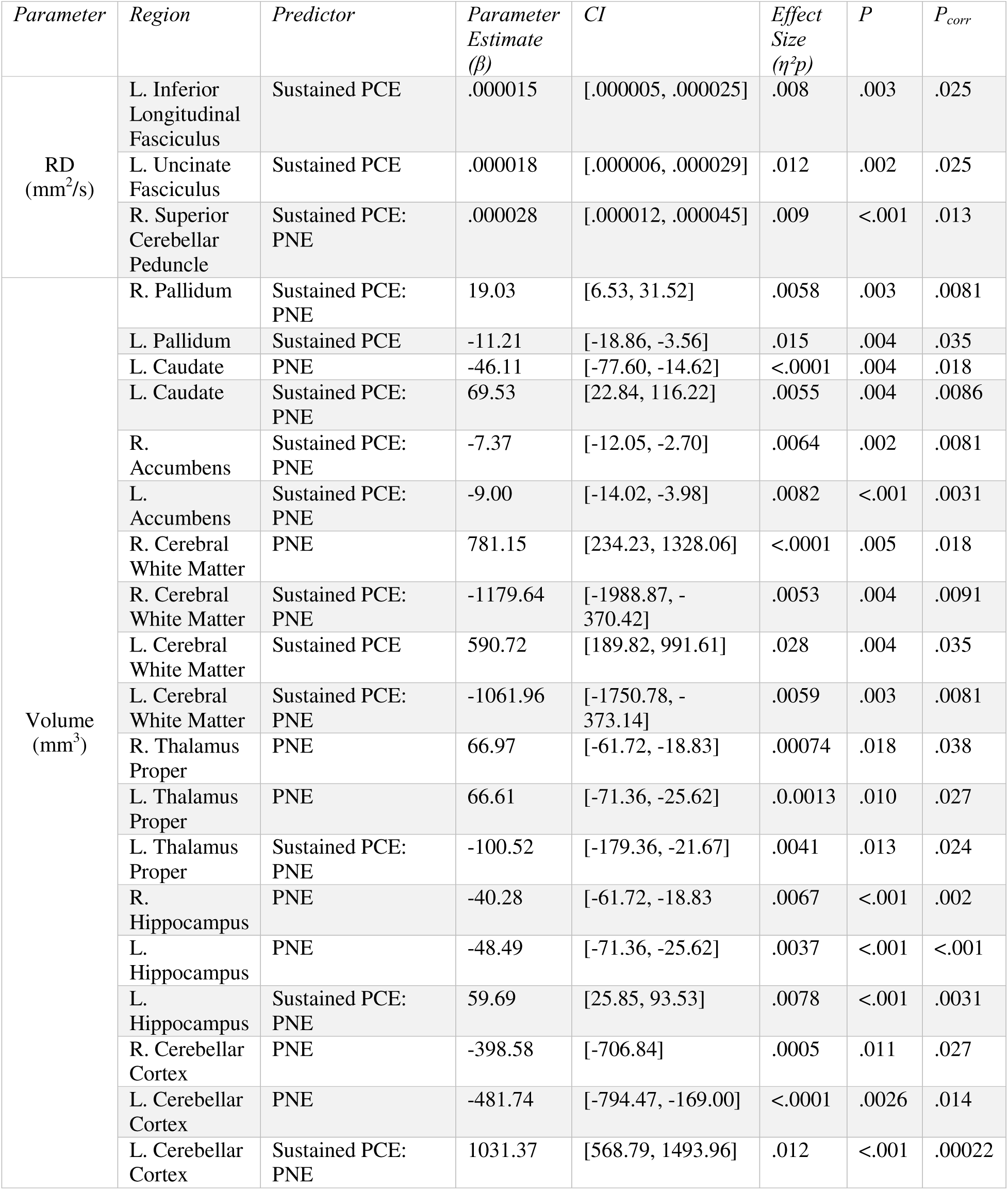
Sensitivity Analysis: Associations Between Sustained PCE and Regional Brain Measures, Covarying for Nicotine Use (PNE) Parameter estimates (β), partial eta squared (η^2^p), andfalse discovery rate-correctedp values (P_corr_) for all statistically significant associations between Sustained PCE, Nicotine use (PNE), and their interaction and regional T2-weighted brain volumes and DTI metrics are presented. Only associations surviving FDR correction (P_corr_ < .05) are shown.

**eFigure 10.**
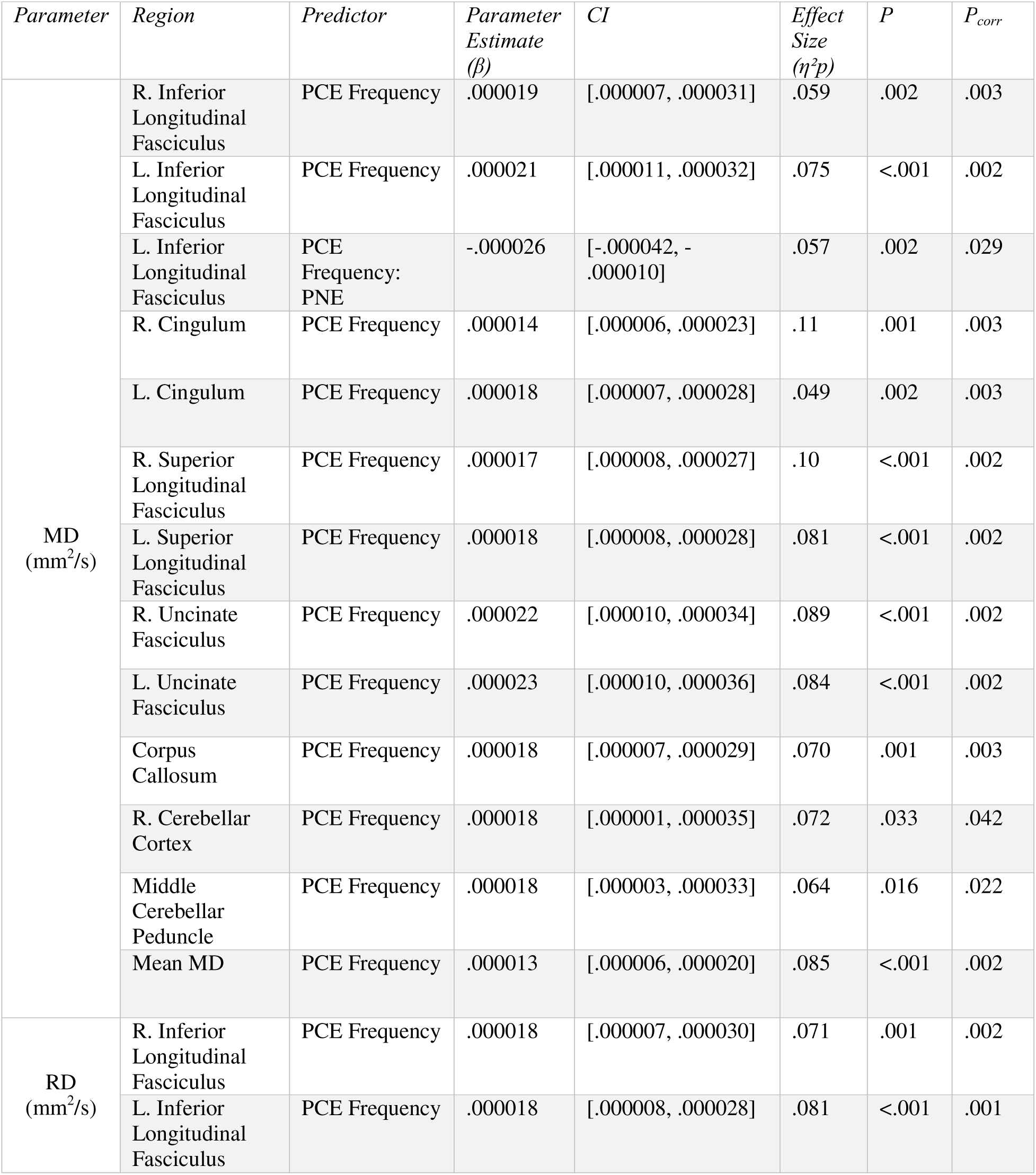
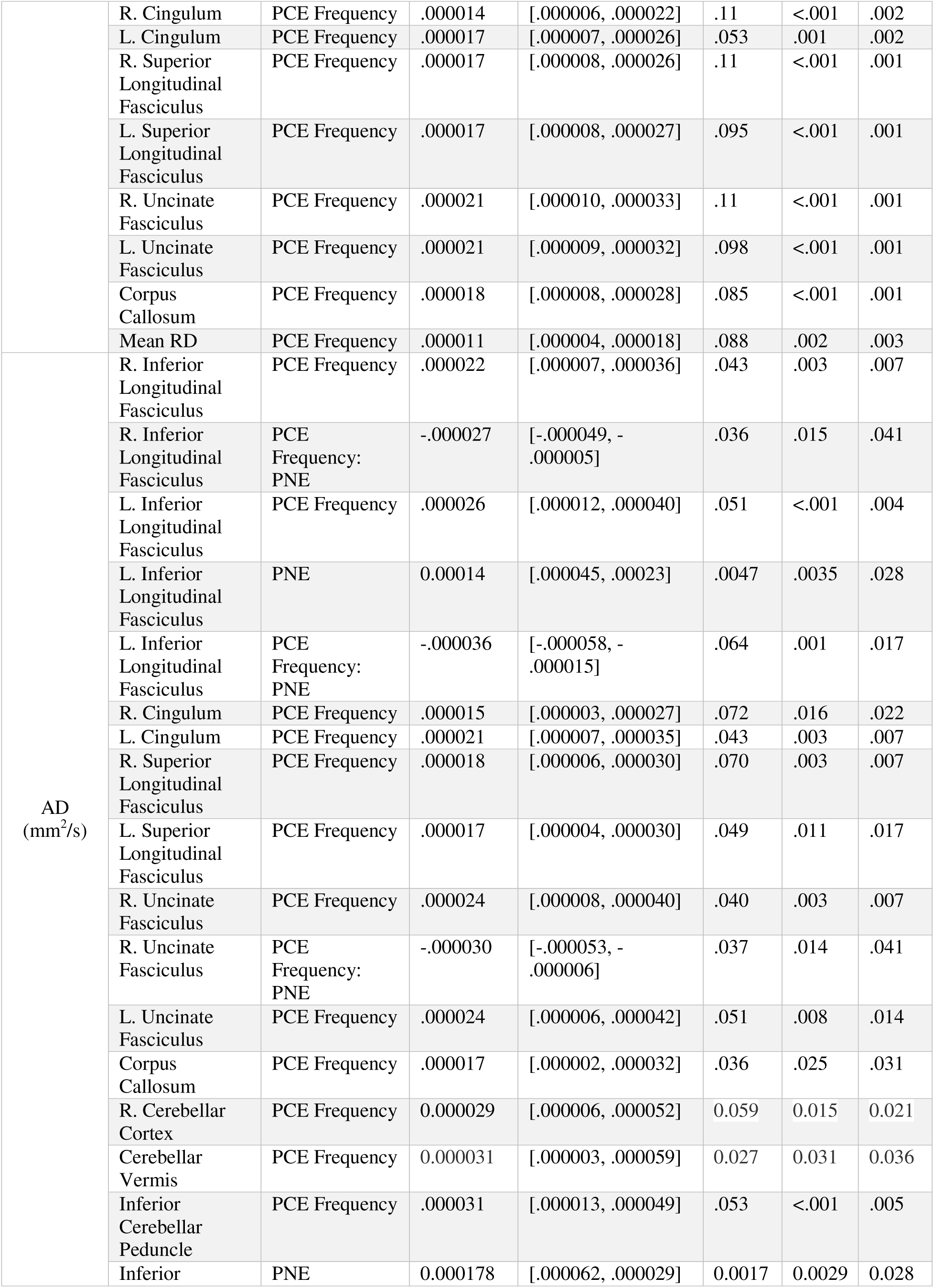
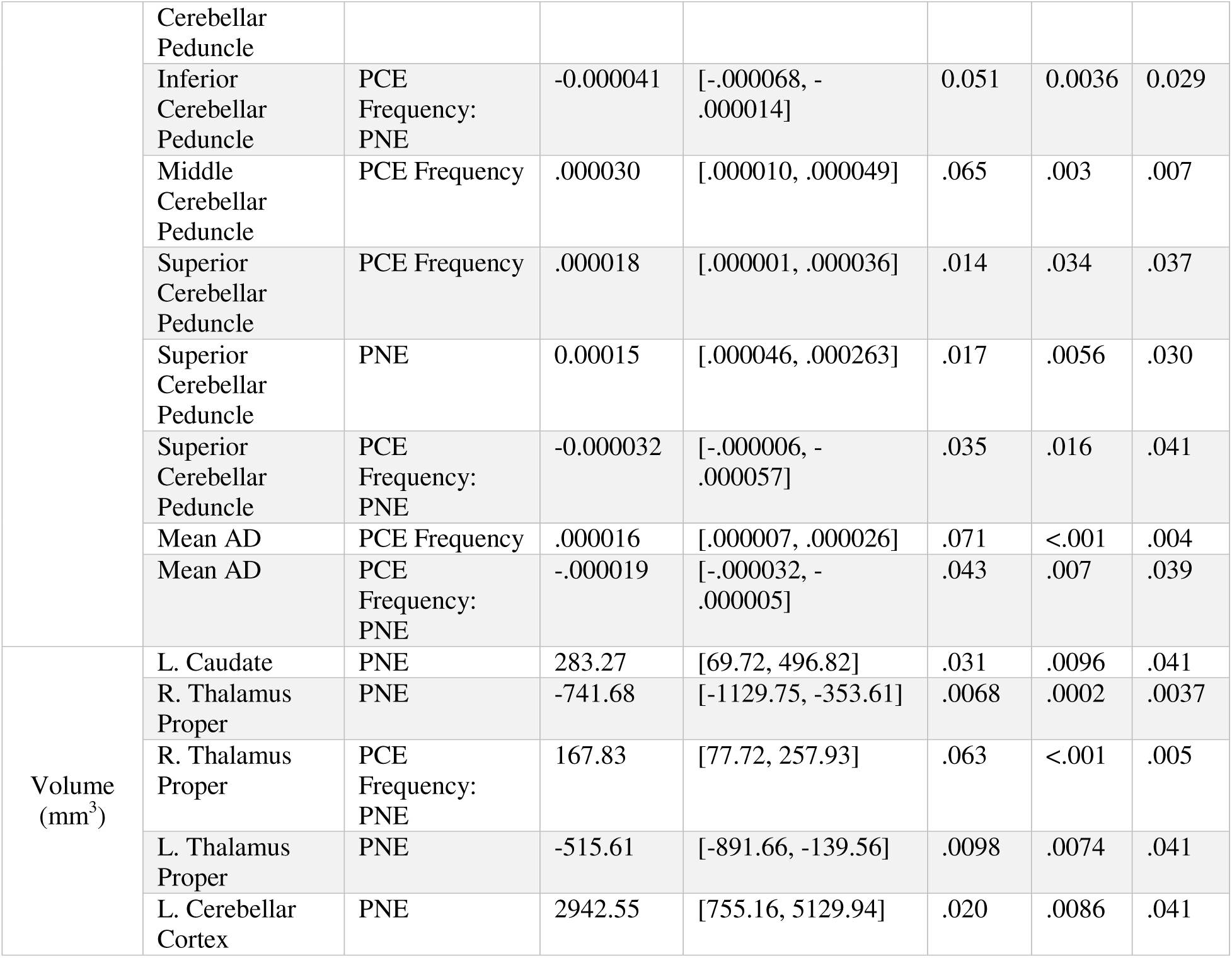
Sensitivity Analysis: Associations Between PCE Frequency and Regional Brain Measures, Covarying for Nicotine Use (PNE) Parameter estimates (β), partial eta squared (η^2^p), andfalse discovery rate-correctedp values (P_corr_) for all statistically significant associations between PCE frequency, Nicotine use (PNE), and their interaction and regional T2-weighted brain volumes and DTI metrics are presented. Only associations surviving FDR correction (P_corr_ < .05) are shown.

