## Supplemental Data for "Associations of Prenatal Cannabis Exposure and Neonatal Brain Development in the HBCD Cohort" (from another version of this preprint: version 3, 2026-07-06)

**eFigure 1. Participant Flowchart.**

*Of 2,310 infants from the HBCD Data Release 2.0, 1,978 had diffusion MRI, 2,072 had volumetric MRI, and 2,114 had complete substance use data. A total of 1,887 participants with complete data (either diffusion MRI or volumetric MRI + complete substance data) were included in the final analytic sample.*

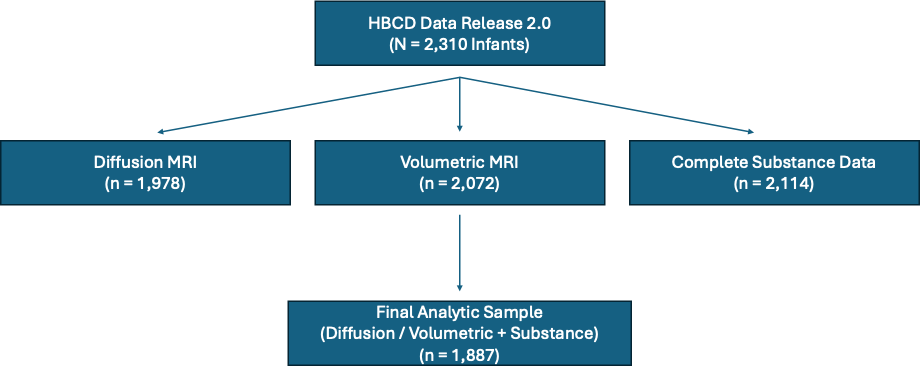

**eFigure 2. Assessment Windows for Cannabis Use Across Pregnancy**

*Cannabis use was retrospectively assessed using the Timeline Follow-Back (TLFB) method for the pre-pregnancy period and for early, mid, and late pregnancy based on timing relative to the last menstrual period and prenatal visits* ^30^. *Blue boxes indicate weeks assessed in PCE timing analyses. The graph indicates the prevalence of reported prenatal cannabis use across TLFB weeks 1-7.*

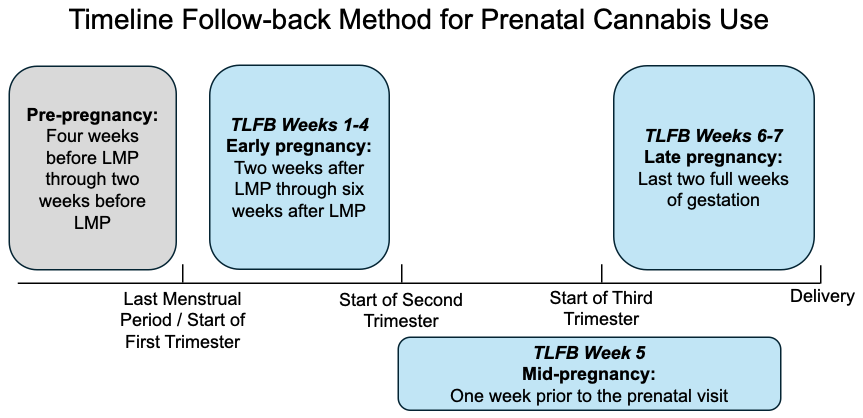

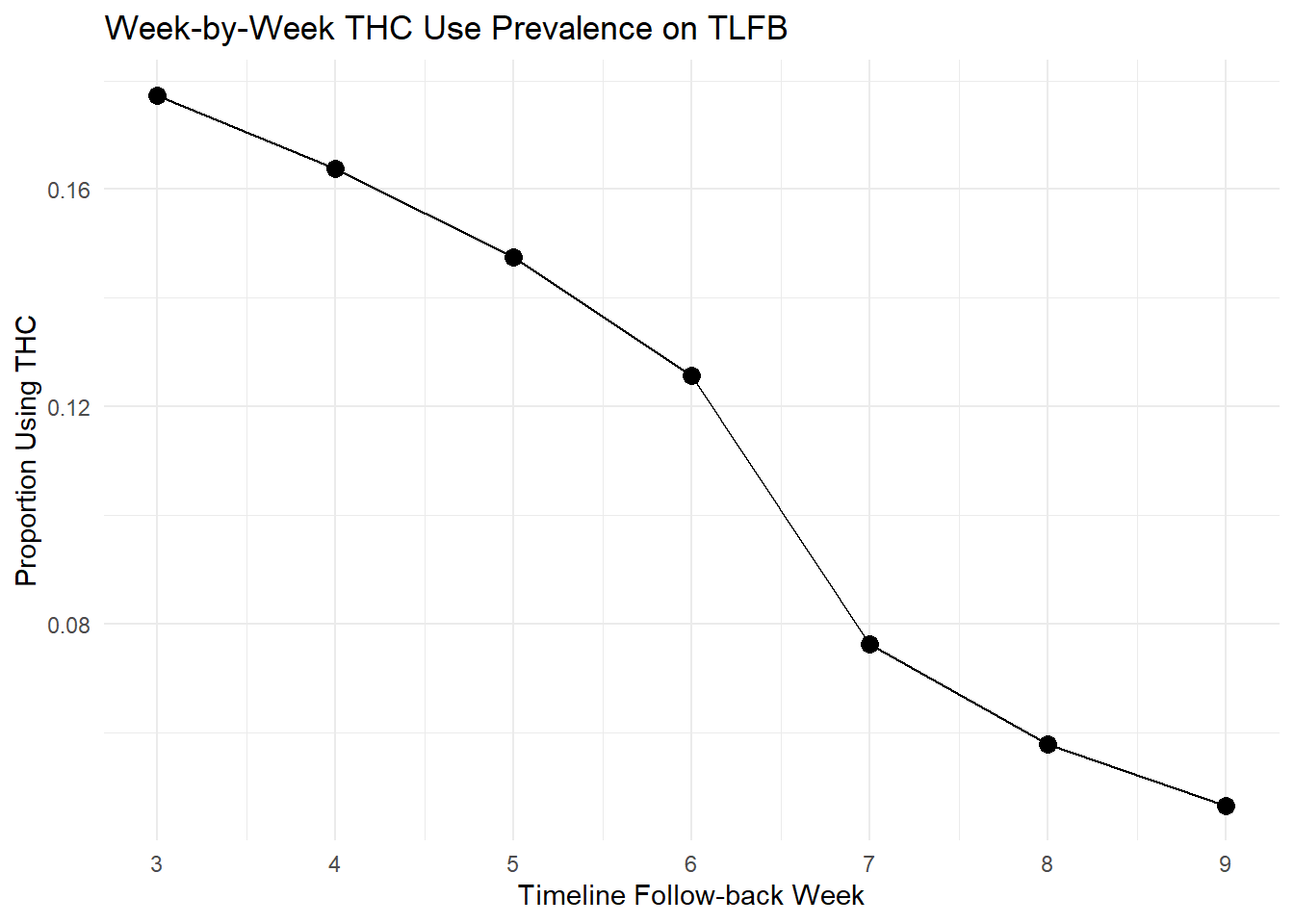

**eFigure 3. Population-Averaged Infant Volumetric Atlas.**

*Sagittal (left), coronal (center), and axial (right) views of the atlas derived from BIBSNet segmentations co-registered to a study-specific ANTs template, visualized in FSLeyes. Regions of interest are labeled.*

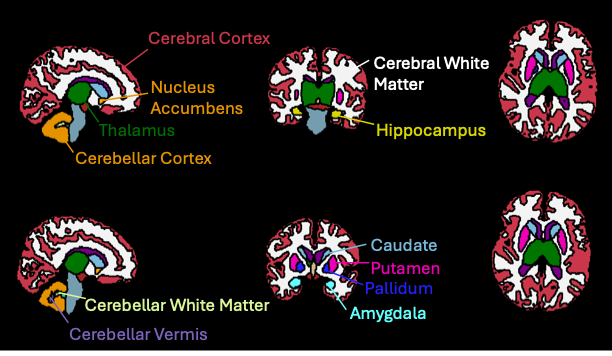

**eFigure 4. Population-Averaged Infant DKI (FA) Atlas.**

*Sagittal (left), coronal (center), and axial (right) views of the atlas derived from dsiPrep tractography co-registered to a study-specific ANTs template, visualized in FSLeyes. Regions of interest are labeled.*

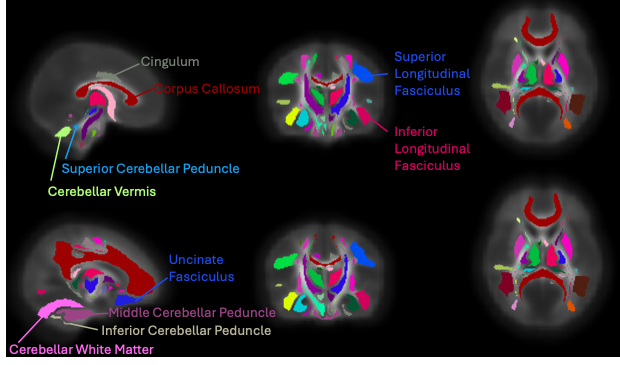

**eFigure 5. Mean Differences in Socioecological Covariates Between Parents With and Without PCE Before and After Covariate Balancing**

*Differences were calculated relative to the unexposed group; negative values indicate lower mean values in the PCE group. Estimates before and after covariate balancing were obtained using the covariate balancing propensity score (CBPS) method* ^41^*. Coral indicates values before balancing; teal, after balancing.*

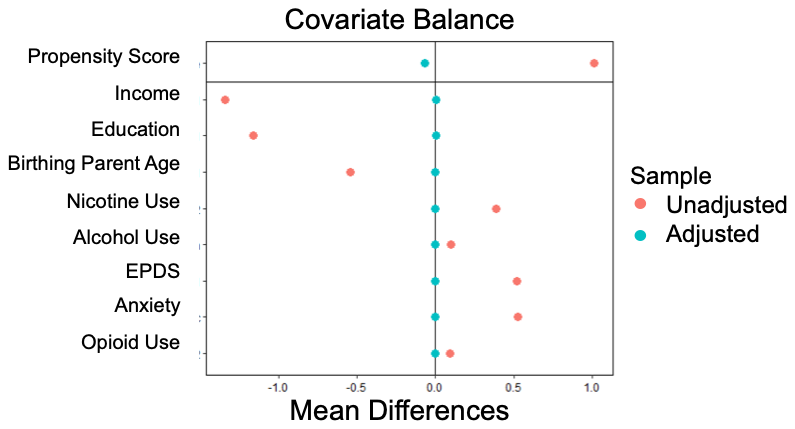

**eFigure 6. Correlations Between Socioecological Covariates and Continuous PCE Before and After Covariate Balancing**

*Correlations are shown for the PCE-only subgroup before and after covariate balancing using the covariate balancing propensity score (CBPS) method* ^41^*. Negative values indicate lower mean values for infants with higher continuous (cumulative) PCE. Coral indicates values before balancing; teal, after balancing. Only predictors that were significantly correlated (p > .05) with PCE frequency were included in the CBPS.*

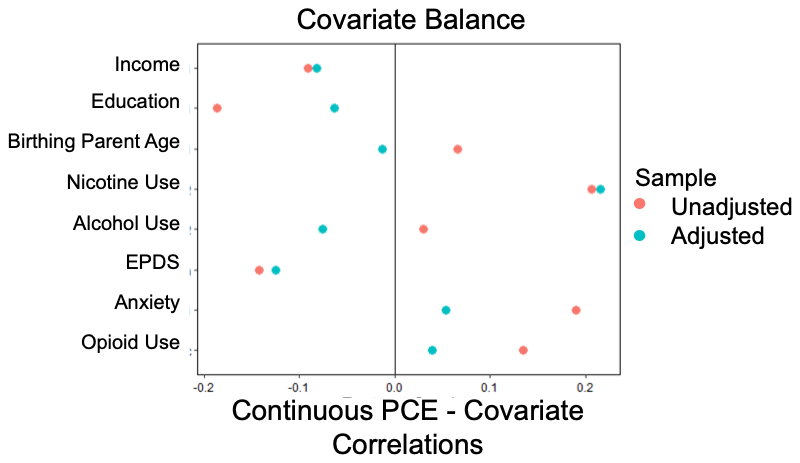

**eFigure 7. Sensitivity Analysis: Associations Between PCE Status and Regional Brain Measures, Covarying for Nicotine Use (NIC)**

*Parameter estimates (ß), partial eta squared (η²p), and false discovery rate–corrected p values (P_corr_) for all statistically significant associations between PCE status, Nicotine use (PNE), and their interaction and regional T2-weighted brain volumes and DTI metrics are presented. Only associations surviving FDR correction (P_corr_ < .05) are shown.*

| *Parameter* | *Region* | *Predictor* | *Parameter Estimate (β)* | *CI* | *Effect Size (η²p)* | *P* | *P_corr_* |
| --- | --- | --- | --- | --- | --- | --- | --- |
| *RD* | L. Association Inferior Longitudinal Fasciculus | PCE | .000014 | [.000006, .000022] | .003 | <.001 | .002 |
|  | L. Association Superior Longitudinal Fasciculus | PCE | .000011 | [.000004, .000018] | .001 | .004 | .011 |
|  | L. Association Uncinate Fasciculus | PCE | .000013 | [.000004, .000022] | .003 | .004 | .011 |
|  | R. Association Uncinate Fasciculus | PCE | .000016 | [.000008, .000025] | .006 | <.001 | .002 |
|  | L. Cerebellar White Matter | PCE | .000021 | [.000010, .000032] | .012 | <.001 | .002 |
|  | Mean RD | PCE | .000008 | [.000002, .000013] | .003 | .009 | .021 |
| *MD* | L. Association Inferior Longitudinal Fasciculus | PCE | .000013 | [.000004, .000022] | .002 | .004 | .015 |
|  | L. Association Superior Longitudinal Fasciculus | PCE | .000012 | [.000004, .000020] | .001 | .004 | .015 |
|  | L. Association Uncinate Fasciculus | PCE | .000015 | [.000006, .000025] | .005 | .001 | .009 |
|  | L. Cerebellar White Matter | PCE | .000021 | [.000009, .000034] | .010 | <.001 | .009 |
| *AD* | L. Association Superior Longitudinal Fasciculus | PCE | .000014 | [.000004, .000025] | .001 | .007 | .036 |
|  | R. Association Uncinate Fasciculus | PCE | .000018 | [.000005, .000031] | .003 | .007 | .036 |
|  | L. Cerebellar White Matter | PCE | .000024 | [.000007, .000041] | .006 | .005 | .036 |
| *Volume* | R. Pallidum | PCE:PNE | 21.196 | [10.941, 31.450] | .009 | <.001 | <.001 |
|  | L. Caudate | PCE:PNE | 59.658 | [21.067, 98.249] | .005 | .002 | .010 |
|  | R. Cerebellar Cortex | PNE | -408.014 | [-694.046, -121.982] | <.001 | .005 | .018 |
|  | R. Cerebellar Cortex | PCE:PNE | 523.892 | [154.508, 893.275] | .004 | .005 | .015 |
|  | L. Cerebellar Cortex | PNE | -500.800 | [-791.594, -210.006] | <.001 | <.001 | .007 |
|  | L. Cerebellar Cortex | PCE:PNE | 796.936 | [421.458, 1172.413] | .010 | <.001 | <.001 |
|  | R. Cerebral White Matter | PCE | 706.068 | [333.186, 1078.950] | .039 | <.001 | .004 |
|  | R. Cerebral White Matter | PNE | 807.340 | [288.297, 1326.383] | .002 | .002 | .013 |
|  | R. Cerebral White Matter | PCE:PNE | -1280.771 | [-1951.066, -610.476] | .008 | <.001 | .001 |
|  | L. Cerebral White Matter | PNE | 627.777 | [178.063, 1077.492] | .012 | .006 | .018 |
|  | R. Hippocampus | PNE | -28.905 | [-49.022, -8.787] | .002 | .005 | .018 |
|  | L. Hippocampus | PNE | -37.640 | [-59.544, -15.735] | .001 | <.001 | .007 |
|  | L. Hippocampus | PCE:PNE | 43.043 | [14.747, 71.339] | .005 | .003 | .010 |

**eFigure 8. Sensitivity Analysis: Associations Between First Trimester PCE and Regional Brain Measures, Covarying for Nicotine Use (PNE)**

*Parameter estimates (ß), partial eta squared (η²p), and false discovery rate–corrected p values (P_corr_) for all statistically significant associations between First Trimester PCE, Nicotine use (PNE), and their interaction and regional T2-weighted brain volumes and DTI metrics are presented. Only associations surviving FDR correction (P_corr_ < .05) are shown.*

| *Parameter* | *Region* | *Predictor* | *Parameter Estimate (β)* | *CI* | *Effect Size (η²p)* | *P* | *Pcorr* |
| --- | --- | --- | --- | --- | --- | --- | --- |
| *RD* | L. Superior Longitudinal Fasciculus | First Trimester PCE:PNE | -.000030 | [-.000046, -.000014] | .013 | <.001 | .002 |
|  | R. Superior Longitudinal Fasciculus | First Trimester PCE:PNE | -.000029 | [-.000044, -.000014] | .013 | <.001 | .002 |
|  | R. Uncinate Fasciculus | First Trimester PCE | .000018 | [.000007, .000029] | <.001 | .001 | .006 |
|  | R. Cerebellar White Matter | First Trimester PCE | .000017 | [.000003, .000031] | .002 | .019 | .040 |
|  | L. Cerebellar White Matter | First Trimester PCE | .000035 | [.000021, .000049] | .006 | <.001 | <.001 |
|  | Vermis | First Trimester PCE | .000022 | [.000007, .000037] | .004 | .004 | .010 |
|  | Middle Cerebellar Peduncle | First Trimester PCE | .000019 | [.000006, .000031] | .007 | .003 | .010 |
|  | Superior Cerebellar Peduncle | First Trimester PCE | .000020 | [.000010, .000029] | .016 | <.001 | <.001 |
|  | Mean RD | First Trimester PCE | .000011 | [.000004, .000018] | .001 | .003 | .010 |
|  | Mean RD | First Trimester PCE:PNE | -.000018 | [-.000030, -.000006] | .010 | .004 | .022 |
| *MD* | L. Superior Longitudinal Fasciculus | First Trimester PCE:PNE | -.000034 | [-.000051, -.000016] | .013 | <.001 | .002 |
|  | R. Superior Longitudinal Fasciculus | First Trimester PCE:PNE | -.000030 | [-.000046, -.000014] | .013 | <.001 | .002 |
|  | L. Cerebellar White Matter | First Trimester PCE | .000036 | [.000020, .000052] | .006 | <.001 | <.001 |
|  | Corpus Callosum | First Trimester PCE:PNE | -.000024 | [-.000043, -.000005] | .005 | .013 | .045 |
|  | Superior Cerebellar Peduncle | First Trimester PCE | .000020 | [.000008, .000032] | .014 | .001 | .005 |
|  | Mean MD | First Trimester PCE:PNE | -.000019 | [-.000033, -.000005] | .009 | .007 | .033 |
| *AD* | L. Superior Longitudinal Fasciculus | First Trimester PCE:PNE | -.000042 | [-.000064, -.000019] | .011 | <.001 | .005 |
|  | R. Superior Longitudinal Fasciculus | First Trimester PCE:PNE | -.000031 | [-.000052, -.000011] | .011 | .003 | .015 |
|  | R. Uncinate Fasciculus | First Trimester PCE | .000023 | [.000006, .000040] | <.001 | .006 | .041 |
|  | L. Cerebellar White Matter | First Trimester PCE | .000042 | [.000020, .000064] | .006 | <.001 | .002 |
|  | Corpus Callosum | First Trimester PCE:PNE | -.000039 | [-.000064, -.000013] | .009 | .003 | .015 |
|  | Superior Cerebellar Peduncle | First Trimester PCE | .000023 | [.000006, .000039] | .010 | .008 | .041 |
| *Volume* | R. Amygdala | First Trimester PCE | -7.284 | [-13.364, -1.204] | .014 | .019 | .046 |
|  | L. Amygdala | First Trimester PCE | -8.371 | [-14.378, -2.364] | .009 | .006 | .034 |
|  | L. Amygdala | First Trimester PCE:PNE | 13.150 | [3.227, 23.072] | .008 | .009 | .026 |
|  | R. Pallidum | First Trimester PCE:PNE | 26.615 | [14.149, 39.081] | .006 | <.001 | <.001 |
|  | L. Caudate | First Trimester PCE | -45.355 | [-73.531, -17.178] | .012 | .002 | .028 |
|  | L. Caudate | First Trimester PCE:PNE | 72.158 | [25.431, 118.884] | .003 | .002 | .011 |
|  | L. Accumbens | First Trimester PCE | -3.536 | [-6.553, -.518] | .001 | .022 | .046 |
|  | L. Accumbens | First Trimester PCE:PNE | 8.160 | [3.135, 13.185] | .017 | .001 | .008 |
|  | R. Cerebellar Cortex | First Trimester PCE | -366.416 | [-636.981, -95.850] | .003 | .008 | .034 |
|  | R. Cerebellar Cortex | PNE | -407.521 | [-693.319, -121.723] | <.001 | .005 | .016 |
|  | R. Cerebellar Cortex | First Trimester PCE:PNE | 668.605 | [219.861, 1117.349] | .003 | .004 | .012 |
|  | L. Cerebellar Cortex | PNE | -503.224 | [-793.508, -212.939] | <.001 | .001 | .006 |
|  | R. Cerebral White Matter | First Trimester PCE | 700.153 | [208.542, 1191.765] | 0.033 | 0.005 | 0.034 |
|  | R. Cerebral White Matter | PNE | 806.350 | [287.062, 1325.639] | 0.002 | 0.002 | 0.013 |
|  | R. Cerebral White Matter | First Trimester PCE:PNE | -1387.650 | [-2203.009, -572.291] | 0.003 | 0.001 | 0.007 |
|  | L. Cerebral White Matter | First Trimester PCE | -505.289 | [-928.003, -82.575] | 0.070 | 0.019 | 0.046 |
|  | L. Cerebral White Matter | PNE | 633.072 | [186.560, 1079.583] | 0.012 | 0.005 | 0.016 |
|  | L. Thalamus Proper | First Trimester PCE | -59.831 | [-107.160, -12.502] | 0.018 | 0.013 | 0.045 |
|  | R. Hippocampus | PNE | -28.876 | [-48.975, -8.776] | 0.002 | 0.005 | 0.016 |
|  | L. Hippocampus | PNE | -37.661 | [-59.539, -15.783] | 0.001 | <0.001 | 0.006 |
|  | L. Hippocampus | First Trimester PCE:PNE | 44.790 | [10.369, 79.212] | 0.001 | 0.011 | 0.026 |

**eFigure 9. Sensitivity Analysis: Associations Between Sustained PCE and Regional Brain Measures, Covarying for Nicotine Use (PNE)**

*Parameter estimates (ß), partial eta squared (η²p), and false discovery rate–corrected p values (P_corr_) for all statistically significant associations between Sustained PCE, Nicotine use (PNE), and their interaction and regional T2-weighted brain volumes and DTI metrics are presented. Only associations surviving FDR correction (P_corr_ < .05) are shown.*

| *Parameter* | *Region* | *Predictor* | *Parameter Estimate (β)* | *CI* | *Effect Size (η²p)* | *P* | *Pcorr* |
| --- | --- | --- | --- | --- | --- | --- | --- |
| *RD* | L. Inferior Longitudinal Fasciculus | Sustained PCE | .000018 | [.000008, .000028] | .011 | <.001 | .004 |
|  | L. Superior Longitudinal Fasciculus | Sustained PCE | .000013 | [.000003, .000022] | .014 | .007 | .032 |
|  | L. Uncinate Fasciculus | Sustained PCE | .000019 | [.000008, .000029] | .014 | <.001 | .005 |
|  | R. Uncinate Fasciculus | Sustained PCE | .000014 | [.000004, .000025] | .010 | .008 | .032 |
|  | Superior Cerebellar Peduncle | Sustained PCE:PNE | .000032 | [.000016, .000048] | .011 | <.001 | .002 |
| *MD* | L. Inferior Longitudinal Fasciculus | Sustained PCE | .000019 | [.000008, .000030] | .011 | <.001 | .012 |
|  | L. Superior Longitudinal Fasciculus | Sustained PCE | .000014 | [.000004, .000024] | .011 | .008 | .036 |
|  | L. Uncinate Fasciculus | Sustained PCE | .000017 | [.000006, .000029] | .013 | .003 | .024 |
|  | Superior Cerebellar Peduncle | Sustained PCE:PNE | .000029 | [.000010, .000049] | .006 | .003 | .041 |
| *Volume* | R. Pallidum | Sustained PCE:PNE | 16.660 | [4.868, 28.451] | .004 | .006 | .016 |
|  | L. Pallidum | Sustained PCE | -10.139 | [-17.416, -2.862] | .014 | .006 | .036 |
|  | R. Accumbens | Sustained PCE:PNE | -7.949 | [-12.382, -3.516] | .007 | <.001 | .002 |
|  | L. Accumbens | Sustained PCE:PNE | -9.199 | [-13.936, -4.462] | .009 | <.001 | .001 |
|  | L. Cerebellar Cortex | Sustained PCE:PNE | 1048.448 | [617.414, 1479.482] | .013 | <.001 | <.001 |
|  | R. Cerebral White Matter | Sustained PCE | 709.405 | [260.298, 1158.512] | .014 | .002 | .017 |
|  | R. Cerebral White Matter | Sustained PCE:PNE | -1188.753 | [-1960.032, -417.474] | .005 | .003 | .009 |
|  | L. Cerebral White Matter | Sustained PCE | 719.077 | [332.912, 1105.243] | .020 | <.001 | .005 |
|  | L. Cerebral White Matter | Sustained PCE:PNE | -1215.120 | [-1878.306, -551.933] | .007 | <.001 | .002 |
|  | L. Hippocampus | Sustained PCE:PNE | 42.617 | [10.122, 75.111] | .004 | .010 | .025 |

**eFigure 10. Sensitivity Analysis: Associations Between PCE Frequency and Regional Brain Measures, Covarying for Nicotine Use (PNE)**

*Parameter estimates (ß), partial eta squared (η²p), and false discovery rate–corrected p values (P_corr_) for all statistically significant associations between PCE frequency, Nicotine use (PNE), and their interaction and regional T2-weighted brain volumes and DTI metrics are presented. Only associations surviving FDR correction (P_corr_ < .05) are shown.*

| *Parameter* | *Region* | *Predictor* | *Parameter Estimate (β)* | *CI* | *Effect Size (η²p)* | *P* | *Pcorr* |
| --- | --- | --- | --- | --- | --- | --- | --- |
| *RD* | L. Inferior Longitudinal Fasciculus | PCE_freq | .000018 | [.000008, .000028] | .070 | <.001 | .001 |
|  | R. Inferior Longitudinal Fasciculus | PCE_freq | .000019 | [.000008, .000030] | .065 | <.001 | .001 |
|  | R. Cingulum | PCE_freq | .000013 | [.000006, .000021] | .093 | <.001 | .001 |
|  | L. Cingulum | PCE_freq | .000017 | [.000007, .000027] | .045 | <.001 | .001 |
|  | L. Superior Longitudinal Fasciculus | PCE_freq | .000017 | [.000008, .000026] | .082 | <.001 | .001 |
|  | R. Superior Longitudinal Fasciculus | PCE_freq | .000017 | [.000008, .000026] | .094 | <.001 | .001 |
|  | L. Uncinate Fasciculus | PCE_freq | .000020 | [.000009, .000031] | .086 | <.001 | .001 |
|  | R. Uncinate Fasciculus | PCE_freq | .000021 | [.000010, .000032] | .097 | <.001 | .001 |
|  | Corpus Callosum | PCE_freq | .000018 | [.000009, .000028] | .074 | <.001 | .001 |
|  | Mean RD | PCE_freq | .000012 | [.000005, .000018] | .075 | <.001 | .001 |
| *MD* | L. Inferior Longitudinal Fasciculus | PCE_freq | .000021 | [.000010, .000031] | .062 | <.001 | .001 |
|  | L. Inferior Longitudinal Fasciculus | PCE_freq:PNE | -.000025 | [-.000041, -.000009] | .055 | .002 | .027 |
|  | R. Cingulum | PCE_freq | .000014 | [.000005, .000022] | .086 | .002 | .003 |
|  | L. Cingulum | PCE_freq | .000018 | [.000007, .000029] | .040 | .001 | .002 |
|  | L. Superior Longitudinal Fasciculus | PCE_freq | .000018 | [.000008, .000028] | .068 | <.001 | .001 |
|  | R. Superior Longitudinal Fasciculus | PCE_freq | .000017 | [.000008, .000026] | .079 | <.001 | .001 |
|  | Uncinate Fasciculus | PCE_freq | .000021 | [.000010, .000032] | .079 | <.001 | .001 |
|  | R. Cerebellar White Matter | PCE_freq | .000020 | [.000003, .000036] | .063 | .022 | .028 |
|  | Corpus Callosum | PCE_freq | .000018 | [.000008, .000029] | .058 | <.001 | .002 |
|  | Middle Cerebellar Peduncle | PCE_freq | .000019 | [.000004, .000034] | .053 | .011 | .015 |
|  | Mean MD | PCE_freq | .000013 | [.000006, .000021] | .070 | <.001 | .001 |
| *AD* | L. Inferior Longitudinal Fasciculus | PCE_freq | .000024 | [.000010, .000038] | .038 | .001 | .005 |
|  | L. Inferior Longitudinal Fasciculus | PCE_freq:PNE | -.000031 | [-.000053, -.000010] | .046 | .005 | .043 |
|  | R. Cingulum | PCE_freq | .000014 | [.000002, .000027] | .053 | .022 | .028 |
|  | L. Cingulum | PCE_freq | .000021 | [.000007, .000035] | .032 | .003 | .008 |
|  | L. Superior Longitudinal Fasciculus | PCE_freq | .000017 | [.000004, .000030] | .038 | .009 | .017 |
|  | R. Superior Longitudinal Fasciculus | PCE_freq | .000017 | [.000006, .000029] | .052 | .004 | .008 |
|  | L. Uncinate Fasciculus | PCE_freq | .000021 | [.000004, .000039] | .034 | .018 | .027 |
|  | R. Uncinate Fasciculus | PCE_freq | .000024 | [.000008, .000039] | .031 | .003 | .008 |
|  | R. Uncinate Fasciculus | PCE_freq:PNE | -.000030 | [-.000053, -.000007] | .037 | .011 | .043 |
|  | Corpus Callosum | PCE_freq | .000017 | [.000003, .000032] | .027 | .021 | .028 |
|  | Middle Cerebellar Peduncle | PCE_freq | .000031 | [.000011, .000050] | .055 | .002 | .008 |
|  | Inferior Cerebellar Peduncle | PCE_freq | .000032 | [.000013, .000051] | .043 | .001 | .005 |
|  | Mean AD | PCE_freq | .000016 | [.000007, .000026] | .055 | <.001 | .005 |
|  | Mean AD | PCE_freq:PNE | -.000019 | [-.000033, -.000005] | .039 | .010 | .043 |
| *Vol* | L. Caudate | PNE | 284.843 | [74.620, 495.065] | .027 | .008 | .036 |
|  | L. Cerebellar Cortex | PNE | 2918.276 | [770.333, 5066.218] | .017 | .008 | .036 |
|  | R. Thalamus Proper | PNE | -716.462 | [-1100.733, -332.191] | .005 | <.001 | .005 |
|  | R. Thalamus Proper | PCE_freq:PNE | 162.712 | [73.833, 251.591] | .059 | <.001 | .007 |
|  | L. Thalamus Proper | PNE | -500.649 | [-871.657, -129.642] | .008 | .008 | .036 |
